# Models, components, and outcomes of palliative and end-of-life care provided to adults living at home: A systematic review of reviews

**DOI:** 10.1101/2025.02.14.25322267

**Authors:** Sophie Pask, Chukwuebuka Okwuosa, Ahmed Mohamed, Rebecca Price, Jennifer Young, Thomas Curtis, Stuart Henderson, Ishbel Winter-Luke, Anisha Sunny, Rachel L. Chambers, Sarah Greenley, Therese Johansson, Anna E. Bone, Stephen Barclay, Irene J. Higginson, Katherine E. Sleeman, Fliss E. M. Murtagh

## Abstract

**Background:** Ageing populations necessitate increased focus on home-based care. The best models and components for community-based palliative and end-of-life care are unknown.

**Aim:** To identify and synthesise review-level evidence on models of palliative and end-of-life care for adults living at home, and examine components of these models and their association with outcomes.

**Design:** A review of narrative, scoping and systematic reviews, using key concepts established *a priori* from Firth et al. and Brereton et al.’s model descriptions. Quality assessment used AMSTAR-2 or equivalent.

**Data sources:** MEDLINE, EMBASE, CINAHL, Cochrane Database, Epistemonikos searched from inception to August 2024, supplemented by CareSearch, PROSPERO, and citation searches.

**Results:** From 6683 initial papers, n=66 reviews were included. Seven models of care were identified; by setting (in-home, outpatient); type of professionals (specialist, integrated, non-specialist); or mode (telehealth, education/training). Components included: holistic person-centred assessment, skilled professionals, access to medicines/care/equipment, patient/family support, advance care planning, integration of services, virtual/remote technology, and education. We categorised outcomes into: i) patient outcomes, ii) family/informal caregiver outcomes, iii) professional outcomes, and iv) service utilisation/cost outcomes. The ‘in-home palliative care’ model was most researched with good evidence of positive benefit. Specialist and integrated models of care were next most researched, with evidence of improved patient and service utilisation outcomes. Cost-effectiveness evidence was lacking.

**Conclusion:** This meta-level evidence supports provision of in-home palliative care, with most review level evidence showing positive effect on patient outcomes. There was also evidence to support specialist palliative care and integration of primary palliative care with specialist support.

**Key statements:** *What is already known about the topic?:*

- Care at home for people approaching the last months or year of life has become increasingly important in recent years, due to the increase in deaths, multimorbidity, and preference of the majority for care at home.
- Individual reviews of the evidence on palliative and end of life care at home have been undertaken, with some evidence of benefit.

*What this paper adds:*

- This paper reports the overall evidence, which largely supports in-home palliative care, especially if delivered via specialist palliative care models or integrated palliative care models (where integration refers to coordination between specialist and non-specialist services).
- It also provides evidence of benefit for education and training, both for informal family carers, and for professionals.
- Detailed narrative synthesis links models of care, with their components and sub-components, and related outcomes.

*Implications for practice, theory or policy:* There is clear evidence supporting provision of in-home palliative care, with common components related to addressing (and delivering positive impact on) patients’ symptoms, psychological distress, and functional status.

## Introduction

Care at home for people approaching the last months or year of life has become increasingly important in recent years, due to the increase in deaths due to a rapidly ageing worldwide population, the increase in multimorbidity, and the priority of the majority for care at home. ^1, 2^ Unplanned and urgent home and community-based care is increasingly also needed and used by those with palliative and end of life care needs. ^3, 4^ Enabling individuals to be cared for and die at home has been identified as a top priority for patients and their family carers, as well as for policy makers. ^5–7^

Palliative care provided at home is delivered by a range of different professionals. This includes those who may provide care day-to-day as part of their wider roles (such as general practitioners/family physicians, community/district nurses, pharmacists, or allied healthcare professionals) and those who specialise in palliative and end of life care (such as specialist palliative care doctors, nurses, allied health professionals, and social workers). ^8^ An essential aspect of enabling patients to stay at home is the care provided around the clock (including care ‘out of hours’; that is between 6pm – 8am, weekends and public holidays), which accounts for over two-thirds of a patient’s week. ^9^ Good quality out-of-hours care has also been identified as an important component of cost-effective care. ^10^ As care provided inside and outside of normal working hours comprises many different service types (including expertise and delivery), there is limited understanding of which service models and components of care work best and for whom, and under what circumstances. ^8, 10, 11^ Evidence is necessary to inform the best type of care needed outside of normal working hours to avoid crises and support people to stay in their preferred place of care, ^5^ and how community care can be a realistic alternative to hospital-based care. ^3, 12^ From a service planning perspective, enabling people to be cared for and die at home in line with their preference is also likely to reduce (proportionately very expensive) hospital admission costs. ^3, 12–14^

Existing reviews of reviews have focused on i) identifying the range of models of palliative care (including care provided at home) ^10, 15^ ii) identifying components of in-home end-of-life care programs and examining their effectiveness (focusing on the last months or days of life), ^11^ iii) quantifying the impact of home palliative care services on whether people will die in a home setting, ^13^ or iv) are focussed solely on out-of-hours services ^16^. However, an over-arching review and synthesis of the evidence on palliative and end-of-life care delivered to adults living at home has not been undertaken.

## Research aim

This review therefore aimed to identify and synthesise review-level evidence on models of palliative and end-of-life care for adults living at home, and examine the components of these models and their association with outcomes.

## Methods

### Study design

A systematic ‘review of reviews’ - useful for combining findings from a large volume of reviews into a single synthesis of evidence ^17^ - and registered on the international prospective register of systematic reviews (PROSPERO Registration Number: CRD42022362156). Reporting adheres to the Preferred Reporting Items for Systematic Reviews and Meta-Analysis (PRISMA) 2020 statement. ^18^ We also adopt the original seven critical items established by Shea and colleagues. ^19^ The main concepts considered as part of this review are defined in ***Box 1***.

##### Box 1. Definitions and considerations around key concepts for this review

**Palliative and end-of-life care** adopts a patient-centred and multidisciplinary approach to providing treatment, care, and support for people with advanced illness, and their families. ^20, 21^ The aim of palliative and end-of-life care is to enable people to have a good quality of life, including symptom management, personal care (such as washing or dressing), support with emotional, spiritual, and psychological needs, and social and family support. ^20, 21^

**Care at home** supports the management of palliative and end-of-life needs in the individual’s home setting (not residential care settings), including enabling a person to remain at home when preferred. This includes care that is provided within ‘normal’ working hours (typically 8am and 6pm from Monday to Friday) and care provided outside of normal working hours (including evenings (generally 6pm until 8am), weekends and public holidays).

**Models of care** are defined as the way in which health and care services are delivered and provides ‘a descriptive picture of practice which adequately represents the real thing’. ^22, 23^ Firth et al established key criteria to define and allow for comparison between models of specialist palliative care, such as the setting of care (e.g. inpatient hospital, inpatient hospice and home-based) or the disciplines delivering care. ^24, 25^ Similarly, Brereton et al focus on structure and multiple components (including who delivers the care, the intervention, setting of care, care recipients, timing and duration, how (e.g. telephone), and purpose (i.e. expected outcomes). ^10^

**Components of care** are defined as attributes, characteristics, features, or elements of a model of care. For example, six essential elements of quality palliative home care have previously been identified, including: integrated teamwork, symptom management, holistic care, skilled providers (who are caring and compassionate), timely and responsive care, and patient and family preparedness. ^26^ We use the term “subcomponent” where elements contribute to single (overall) care component.

**Outcomes** are defined here as ‘the change in a patient’s current and future health status attributed to preceding healthcare’. ^27^ For this systematic review, we consider outcomes in relation to models and components of care and endeavour to reflect how the models and components relate to outcomes of care. We report outcomes as: patient outcomes, family or informal caregiver outcomes, professional outcomes, and service utilisation and costs.

### Eligibility criteria

We included reviews which reported evidence on home or community-based palliative and end-of-life care. Detailed inclusion and exclusion criteria are shown in ***Table 1***.

**Table 1.** Inclusion and exclusion criteria.

| Inclusion | Exclusion |
| --- | --- |
| *Population:* Adults (i.e. aged $\geq 18$) living at home with palliative and end-of-life care needs and their informal caregivers. Reviews considering various age ranges provided the focus of the research is adults. | Reviews solely considering children and adolescents (i.e. aged $\leq 18$). |
| *Exposure:* Treatment, care, and/or support provided to adults living at home with palliative and end-of-life care needs within the community setting. There is a clear discussion regarding treatment, care, and/or support provided to those living at home where multiple settings are discussed. | Focuses on care provided in hospital or other institutional settings (such as inpatient hospice, care home, or other residential settings), or transitions between settings of care. There is no clear discussion regarding treatment, care, and/or support provided to those living at home where multiple settings are discussed. |
| *Outcomes:* All outcomes related to patients, | No reviews will be excluded based on the |
| families, services, and costs will be examined. | outcomes reported. |
| *Study type:* Review-level evidence that explicitly and systematically reports on all types of original study (e.g. intervention, observational and qualitative studies). | Publications that are not review-level evidence (i.e. primary studies). Reviews that have not used explicit and systematic methods to collate and synthesise findings (i.e. lacks a clearly formulated research question or information about the evidence search, selection methods, analysis, and synthesis). Opinion papers, editorials, and conference abstracts. |
| *Language:* Written in English. | Not written in English. |

### Search strategy

The search strategy was developed with an information specialist (SG) drawing on relevant reviews to refine search terms. The search strategy was refined using ‘sentinel’ reviews (reviews we expected to find through searching). Hence, scoping searches were used to refine our approach; for example, initial searches revealed that although most of the gold standard papers were identified, some reviews that report on out-of-hours palliative and end-of-life care within a primary care context were not identified. Therefore, final searches used database appropriate subject headings terms and keywords relating to palliative care, primary care, home-based or community setting, and out-of-hours (see Supplemental material 1 for all search strategies for databases used), combined with study design filters for reviews.

MEDLINE, EMBASE (via OVID), CINAHL (via EBSCOHost), Cochrane Databases of Systematic Reviews (via Cochrane library) and Epistemonikos (via www.epistemonikos.org) were originally searched from inception to September 2021, with updates in December 2022, and September 2023. We also searched the CareSearch Project systematic review collections which consolidates online palliative care knowledge ^28^ and the following systematic review collection subjects were reviewed and cross-checked for missing reviews: site of care, models of service delivery, place of death, general practitioners and occupational therapists. In addition, PROSPERO was used as a source to check for any additional reviews. The date of the last search for all sources was August 1 2024. Forward and backward citation searching of included reviews using Citation Chaser was used to identify any additional reviews that potentially meet the inclusion criteria. Returned records from the search were imported into EndNote 21 ^29^ for deduplication, and then transferred to another reference management software; Rayyan. ^30^ This software allowed for remote working and collaboration within a distributed team.

### Review selection

The titles and abstracts of retrieved results were independently screened by at least two review authors (SP, RP, CO, AM, and AS) against the inclusion criteria. The full texts of potentially eligible reviews were then retrieved and independently assessed for eligibility by two review authors (SP, RP, CO, AM, and AS). Where there was any disagreement between the two review authors over the eligibility, this was resolved through discussion with a third review author (FM).

### Data extraction

The development of the data extraction template was guided by the definitions provided by Firth et al. ^24, 25^ and Brereton et al., ^10^ and considering the descriptive picture of practice that was likely to be provided by the authors of reviews (see ***Box 1***). The development of the data extraction form was an iterative process that considered the models and components of care described and reported within each review, including: target population of the review (i.e. care recipients), setting of care, health and social care professionals involved in delivering care (including presence of multiple disciplines), the intervention, and objectives of care (including mode of delivery, and timing and duration), outcomes reported in the review, and the clinical effectiveness and cost-effectiveness of the interventions (i.e. models of care and components). Extracted information was managed using Microsoft Excel (2019). ^31^ Five review authors (SP, RP, CO, AM, and AS) independently completed data extraction from the included reviews, including the models of care, components, and outcomes. 10% were double extracted and compared at the start of extraction to help standardise extraction. Any discrepancies identified were resolved through discussion.

### Quality assessment

The quality of included reviews was assessed independently by at least two review authors (TC, AM, JY, and CO) depending on the methodology of the included review using one of the assessment tools:

For reviews which were systematic (i.e. systematic reviews, rapid systematic reviews, umbrella reviews conducted systematically, and integrative reviews conducted systematically), we used A Measurement Tool to Assess Systematic Reviews (AMSTAR-2). ^19^ This is a 16-component critical appraisal tool for systematic reviews that include randomised and non-randomised studies of healthcare interventions that generates a descriptive judgement of a review’s quality by evaluating critical and non-critical domains, has been recommended for its precision, critical domains, risk of bias assessment, and wide acceptability. ^32–34^ We classified the quality appraisal results from AMSTAR-2 as critically low in quality, low in quality, moderate, or high in quality, depending on the severity of the flaws. Critically low quality indicates multiple flaws with or without weaknesses; low quality indicates one flaw with or without non-critical weaknesses; moderate indicates more than one non-critical weakness but no flaw; and high quality indicates no or one non-critical weakness without flaw. Pieper and colleagues demonstrated that assessments of which domains are considered critical can vary, which can impact on the overall evaluation of a review’s quality – using AMSTAR-2. ^35^For narrative reviews that followed a systematic approach, we used the Scale for the Assessment of Narrative Review Articles (SANRA). ^36^ SANRA, a brief critical appraisal tool for the assessment of non-systematic articles has good specificity for narrative reviews, internal consistency, and inter-rater reliability for multiple reviewers. ^36^ The six items that form the revised SANRA scale are rated in integers from 0 (low standard) to 2 (high standard), with 1 as an intermediate score. The maximal sum score is 12. ^36^For scoping reviews that followed a systematic approach, we assessed them based on their adherence or not, to the Preferred Reporting Items for Systematic Reviews and Meta-analyses Extension for Scoping Reviews (PRISMA-ScR) checklist ^37^ or the Levac et al methodology for scoping reviews. ^38^No reviews were excluded from the synthesis due to poor quality. However, the quality assessments were used to inform critical reflection on the strengths and limitations regarding the robustness of the synthesis.

### Data Synthesis

The synthesis aimed to explain the models and components of care identified, whether any components identified could be related to evidence of effectiveness (i.e. any significant positive effects on any of the outcomes measured) or cost-effectiveness.

Due to expected heterogeneity in the included reviews, we conducted a narrative synthesis following the guidance outlined by Popay and colleagues. ^39^ The initial development of the data extraction form involved identifying models, components, and outcomes within and across the reviews. Hence, we developed a preliminary synthesis to summarise and organise findings. Where reported, the definitions and descriptions of the reported models and components of care were summarised. The evidence reported in each review was tabulated using textual descriptions to provide a descriptive summary of the elements earlier described and explore relationships. The relationships between models and components, then models and outcomes were explored and summarised (e.g., chart/matrix) by quantifying the reviews that explored the identified model, component, or outcome, and then developing an accompanying narrative synthesis. Applying exploratory subgroup analysis, we aimed to understand the differences in the components and outcomes and group these as subgroups.

## Results

### Results of review selection process

Six thousand, six hundred and eighty-three records were identified from the searches. Duplicates (n = 2473) were removed. Following title and abstract screening (n = 3963), n = 473 records were retrieved for full text screening. Sixty-six reviews were eligible for inclusion. Full details are shown in a PRISMA flow diagram ^18^ (***Figure 1***).

**Figure 1:**
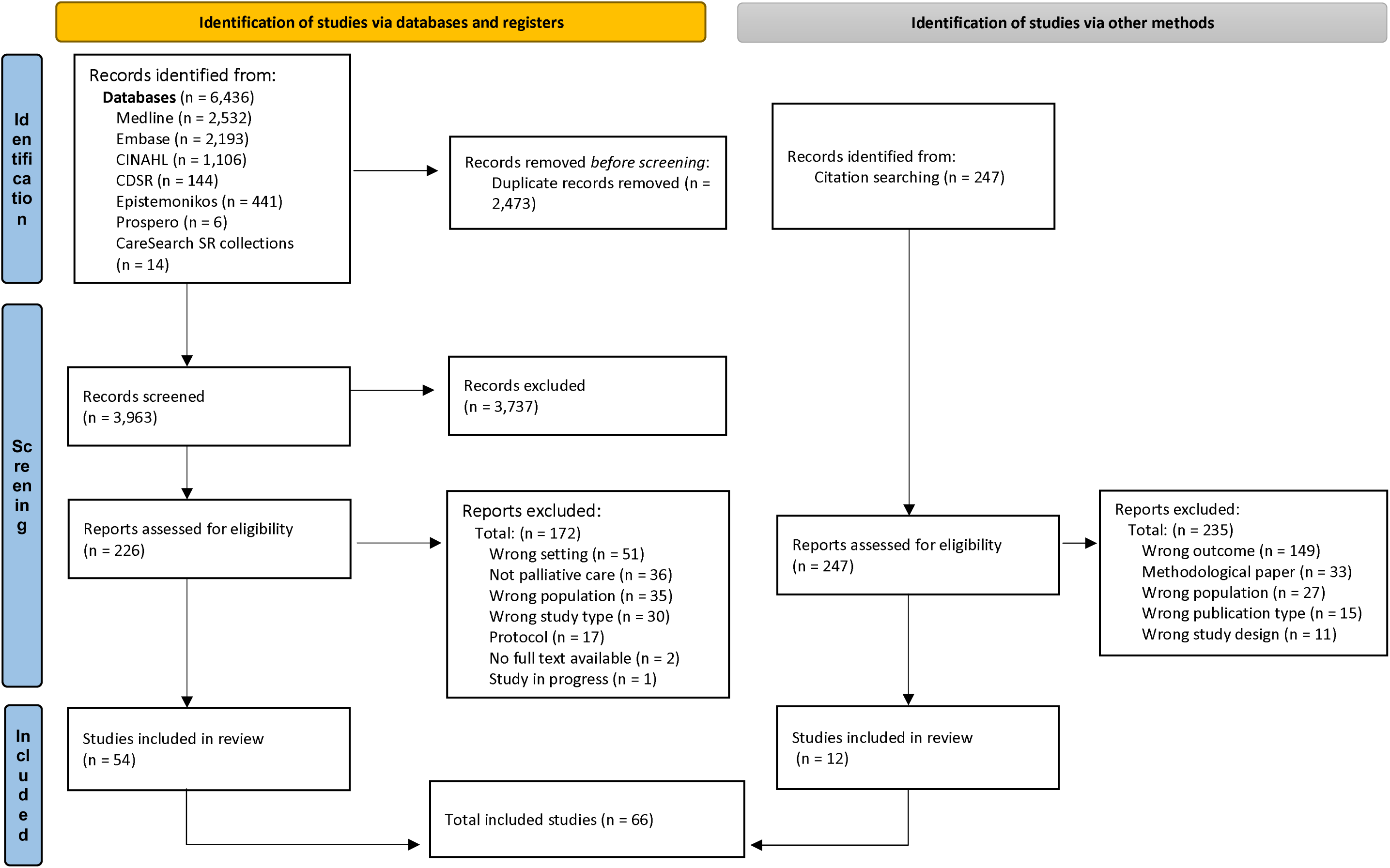
PRISMA flowchart ^18^

### Description of included reviews

The included reviews (n = 66) had varying publication dates with the majority published from 2019 onwards (n = 47). ^16, 24, 40–84^ Fourteen reviews were published between 2013 – 2018 ^10, 11, 13, 15, 85–94^ two between 2007 – 2012 ^95, 96^ and three prior to 2007. ^97–99^

The reviews were conducted across various regions, with the majority in Europe (n = 25), ^10, 13, 16, 41, 42, 45, 48, 50, 51, 53, 54, 56, 60, 70–73, 78, 80, 81, 90, 95, 96, 98, 99^ North America (n = 18), ^11, 40, 46, 49, 52, 55, 58, 63, 67, 69, 82, 84, 87–89, 91, 94, 97^ Australia (n = 7), ^15, 47, 64, 66, 85, 86, 93^ Asia (n = 2), ^44, 62^ South America (n = 2), ^75, 77^ and Africa (n=1).^76^ Reviews were also published from more than one continent (n = 11). ^24, 43, 57, 59, 61, 65, 68, 74, 79, 83, 92^

The study design of the included reviews was categorised for consistency into reviews which we judged were systematic (n = 51), ^10, 11, 13, 15, 16, 40, 42–47, 49–53, 55–64, 66, 68, 71, 74–82, 84, 86–88, 90, 92–94, 96–99^ narrative reviews (n = 6), ^24, 54, 65, 85, 91, 95^ and scoping reviews (n = 9). ^41, 48, 67, 69, 70, 72, 73, 83, 89^ The sources of data in the included reviews included experimental, quasi-experimental, feasibility studies, economic evaluations, cohort and observational studies, case studies, qualitative studies, and mixed methods studies.

The populations and comparators included in the reviews varied; however, most were concentrated on patients and caregivers, with ‘usual care’ provided for the comparator group. Further details of these reviews can be found in Supplemental material 2: Characteristics of Included Reviews.

### Results of quality appraisal

Fifty-two of the included reviews conducted a quality appraisal of their included studies using different tools while fourteen did not conduct any quality appraisal. Using AMSTAR-2, ^19^ we appraised fifty-two reviews as follows: Critically low quality (n = 28) ^10, 11, 15, 40, 43, 46, 49, 50, 55–59, 62, 63, 71, 77, 81, 84, 87, 88, 90, 93, 94, 96–99^ low quality (n = 14) ^16, 42, 45, 52, 53, 60, 64, 66, 68, 75, 78, 82, 86, 92^ moderate quality (n = 5), ^44, 47, 51, 74, 76^ and high quality (n = 4). ^13, 61, 79, 80^ One scoping review was reported according to PRISMA guidelines, thus assessed with AMSTAR-2. ^89^ Using SANRA, ^36^ we appraised six narrative reviews as 9/12 (n = 2), ^91, 95^ 11/12 (n = 1), ^85^ and 12/12 (n = 3) ^24, 54, 65^ (higher scores represent better quality; categorisation e.g. to low/medium/high categories is not recommended). Six scoping reviews adhered to the PRISMA-ScR ^37^ checklist, ^41, 48, 67, 72, 73, 83^ while one did not adhere to PRISMA-ScR or any equivalent, ^69^ and one utilised Levac et al., ^38^ methodology for scoping reviews instead of PRISMA-ScR checklist. ^70^ We did not exclude any review based on review quality. Further details can be found in Supplemental material 2.

### Models of care

We identified seven models of care from the included reviews, including ‘education and training’ as a ‘model of care’ for clarity, while recognising education and training is only indirectly (rather than directly) related to care. We categorized these models according to: setting of care; professional group delivering the care; mode of delivery of care; and education and training to support delivery of care (See ***Table 2***). A number of reviews reported on multiple models of care.

**Table 2:**
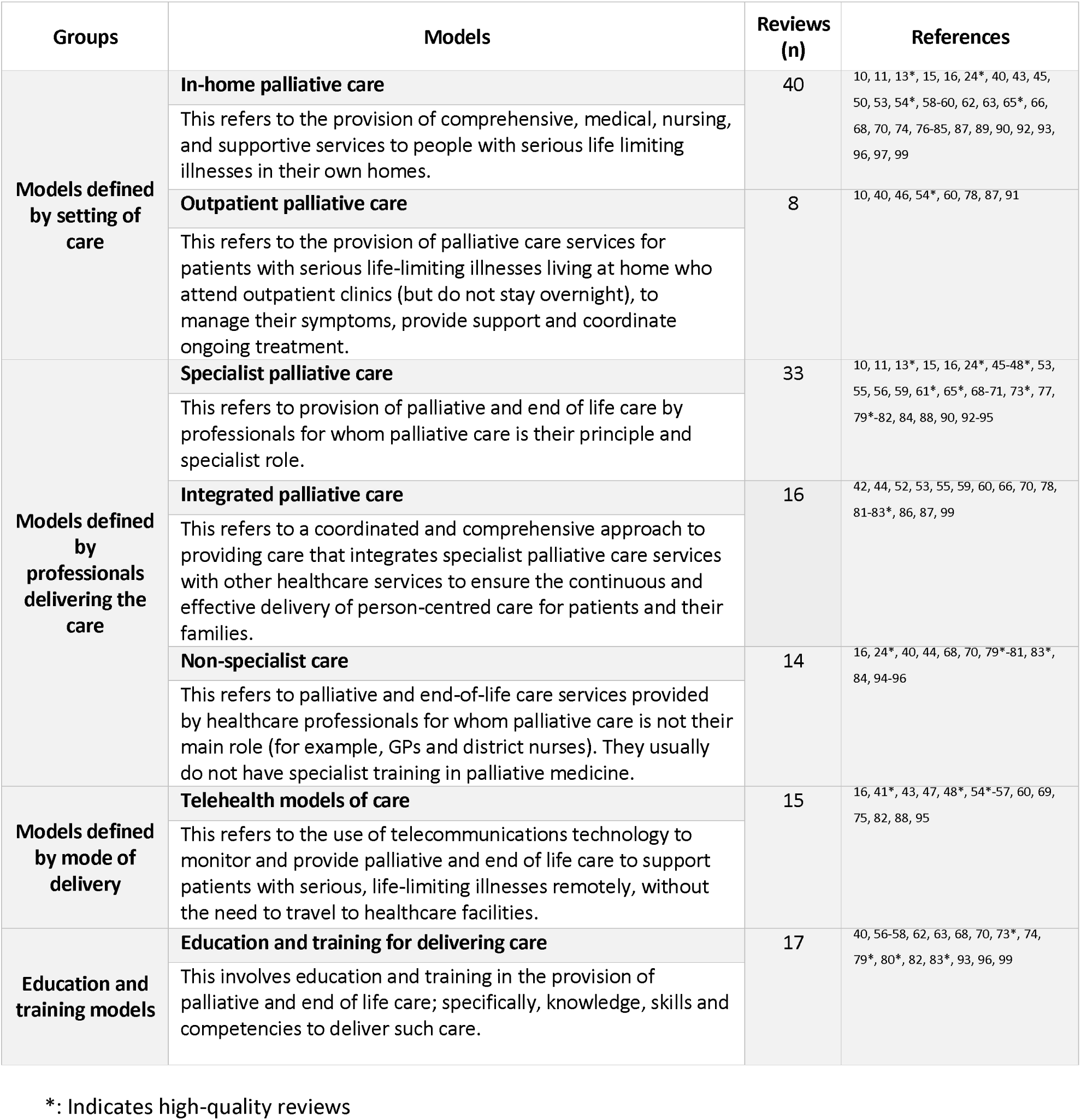
Identified models of care.

The In-home model had the most evidence (n = 40 sources) while the outpatient model had the least (n = 8 sources).

### Components and subcomponents of care

Within the seven models of care, we identified components and subcomponents. These components and subcomponents were derived from 89.4% (n = 59) of included reviews (seven reviews did not describe components or subcomponents). The main components (often common across the seven models) were: a) holistic and person-centred assessment (n = 35, 53.0% of included reviews), b) skilled professional care (n = 40, 60.6% of included reviews), c) access to medicines, care and equipment (n = 16, 24.2% of included reviews), d) support for patients and their families (n = 43, 65.2% of included reviews), e) advance care planning (n = 27, 40.9% of included reviews), f) integration of services (n = 31, 47.0% of included reviews), g) virtual and remote technology (n = 19, 28.8% of included reviews), and h) educational interventions (n = 32, 48.5% of included reviews) (see ***Table 3***).

**Table 3:**
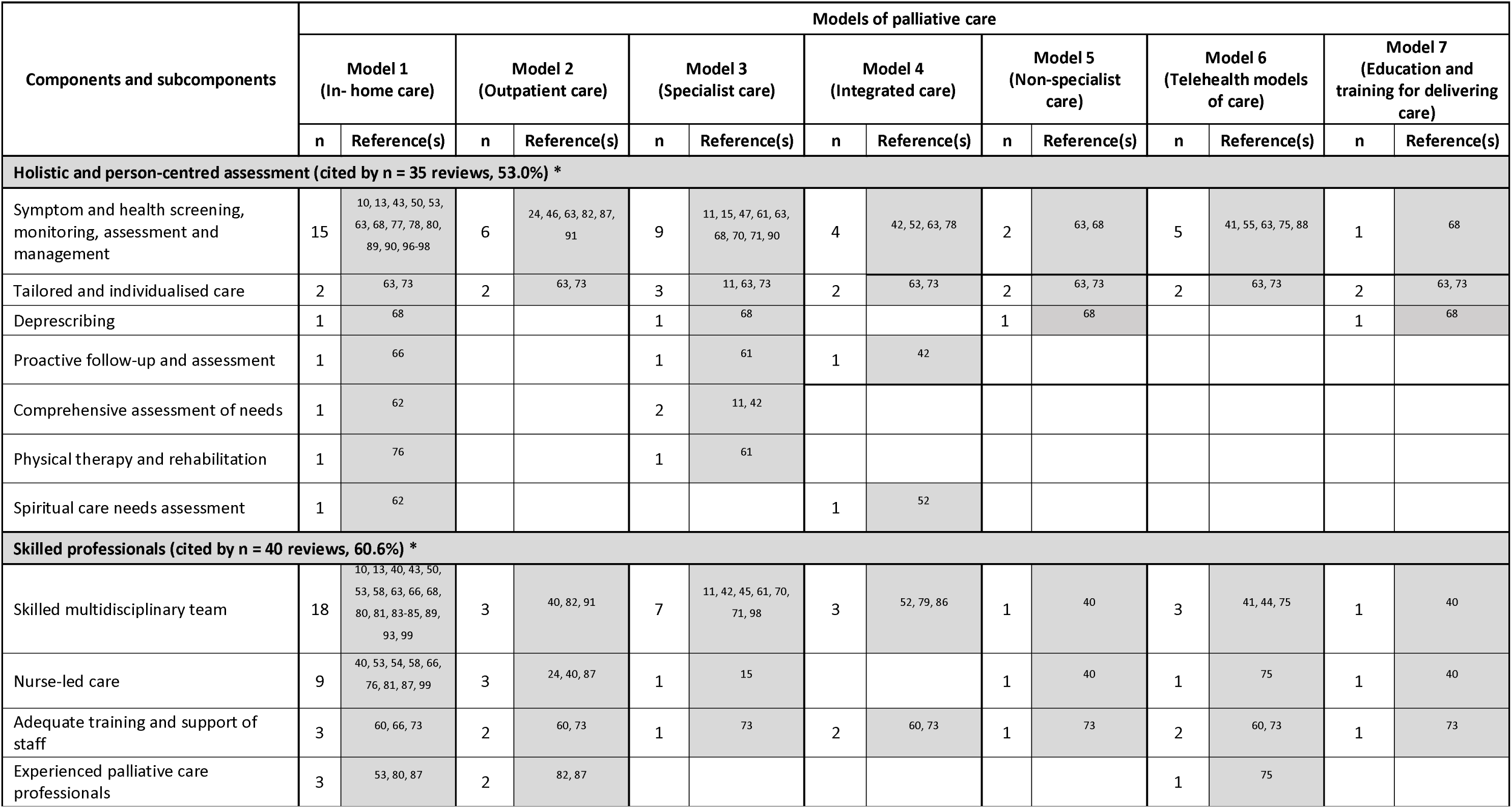
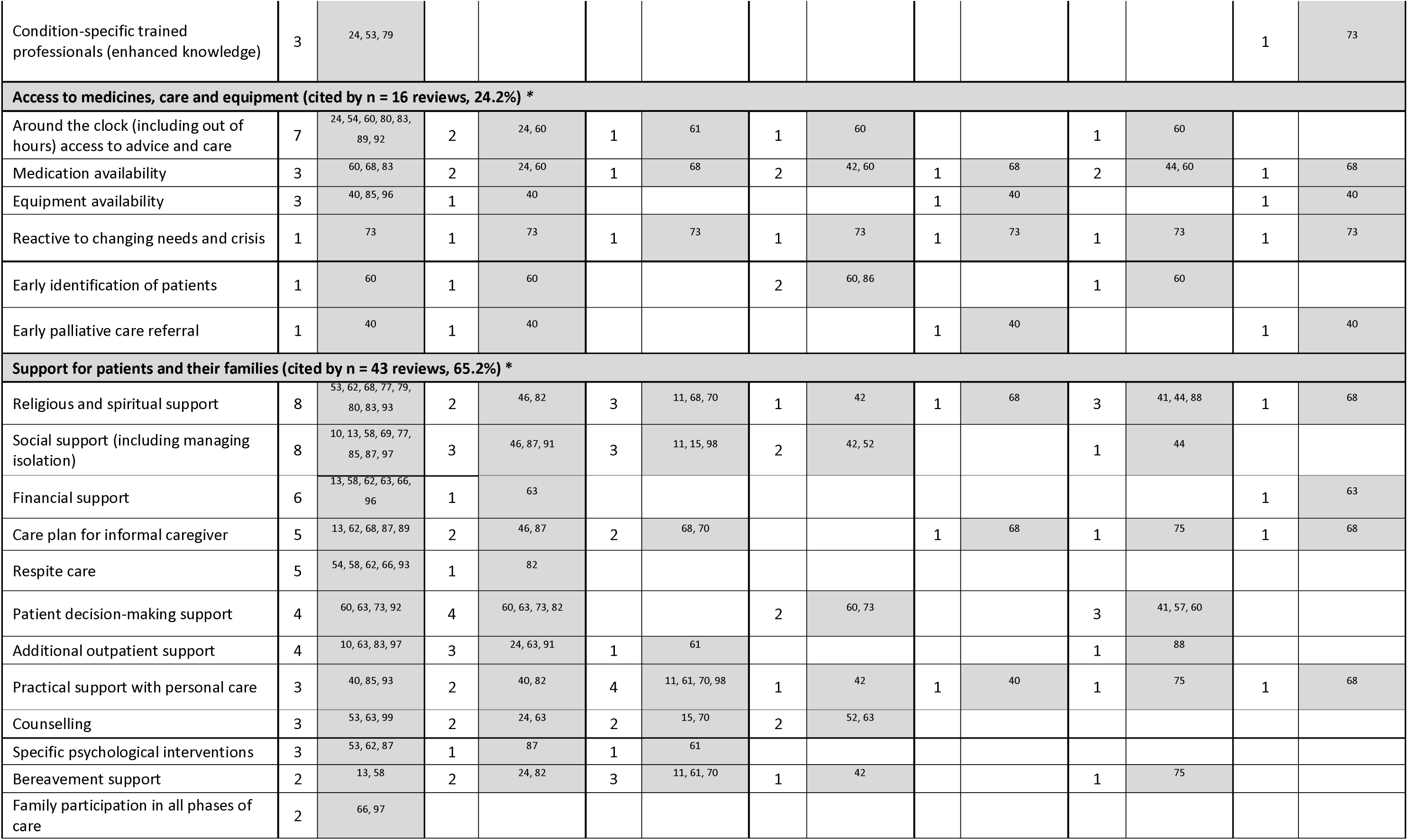
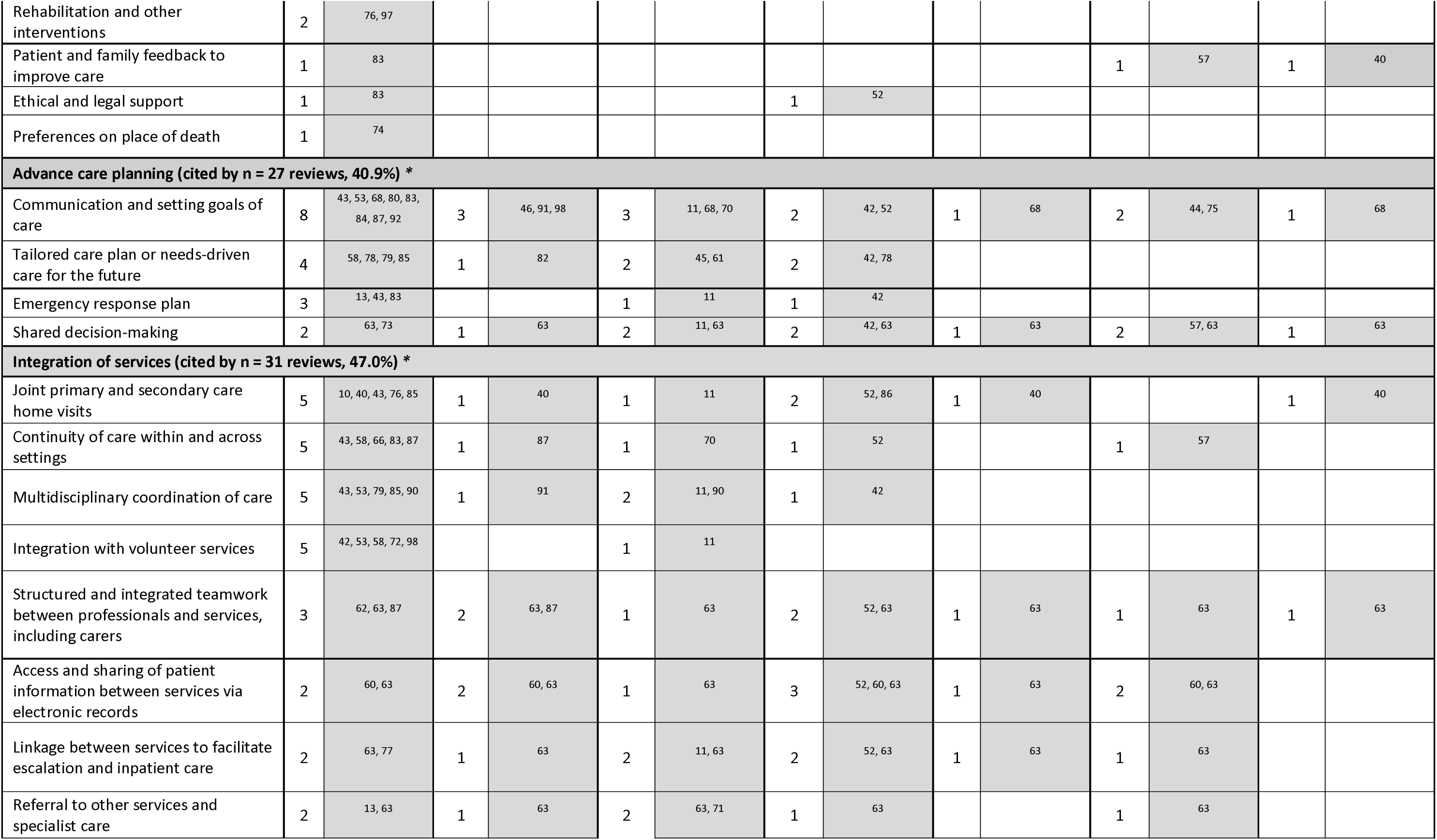
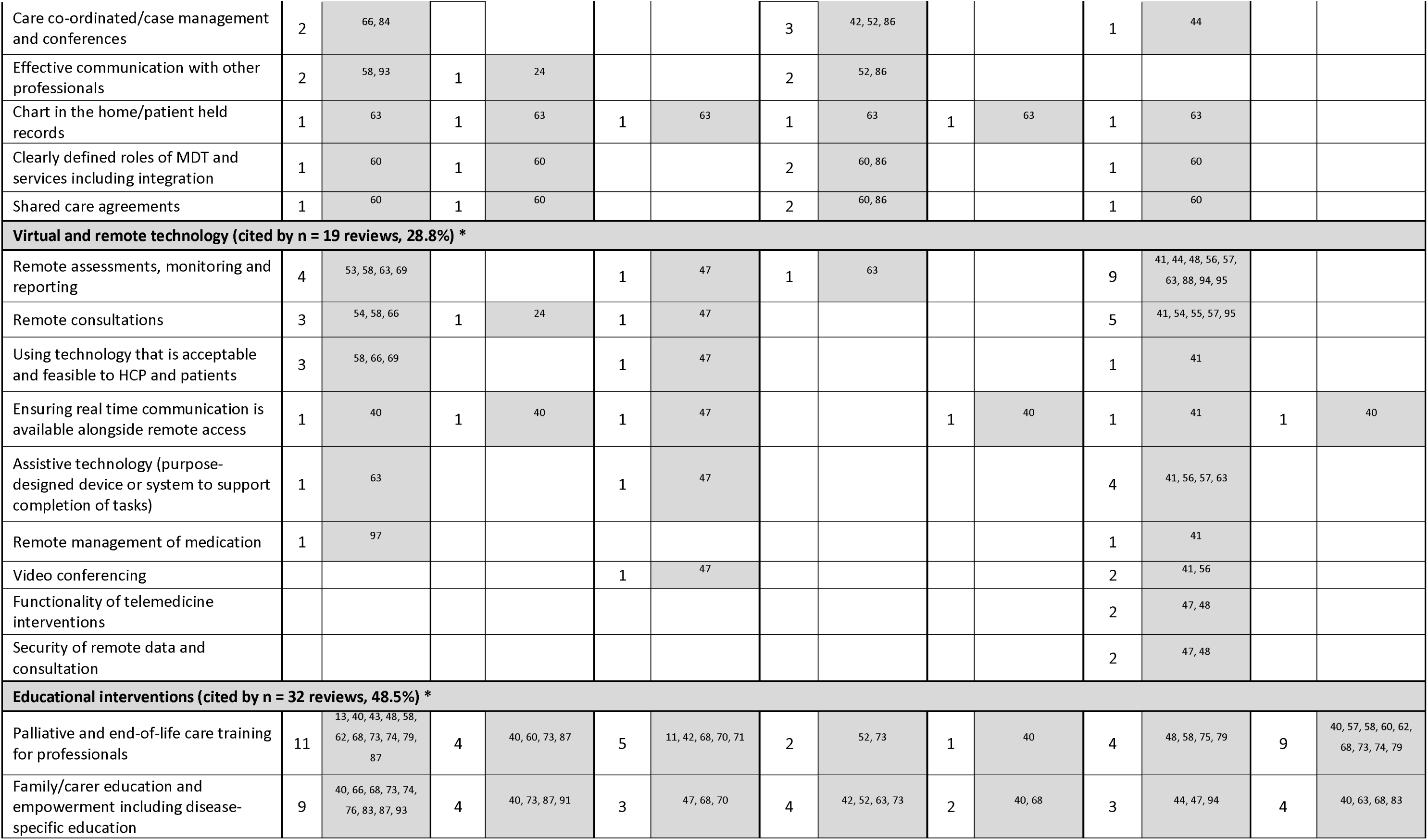
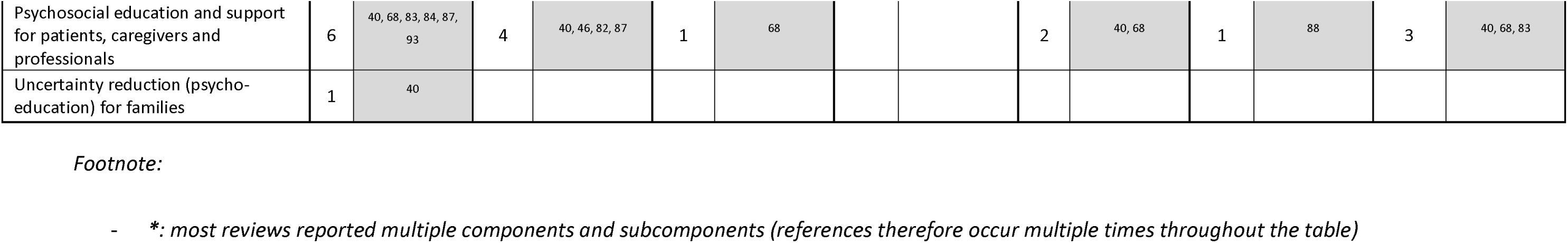
Numbers of included reviews reporting on the components and subcomponents of care, grouped by the seven different models of care we identified.

Sixty-four subcomponents of care were identified and grouped under the appropriate component above. The most frequently reported subcomponents were ‘skilled multidisciplinary team’ (n = 33, 50.0% of included reviews), under the component ‘skilled professionals’, and ‘symptom and health screening, monitoring, assessment and management’ (n = 31, 47.0% of included reviews), under the component ‘holistic and person-centred assessment’. See ***Figure 2*** for details of the components, grouped by the identified models of care.

**Figure 2:**
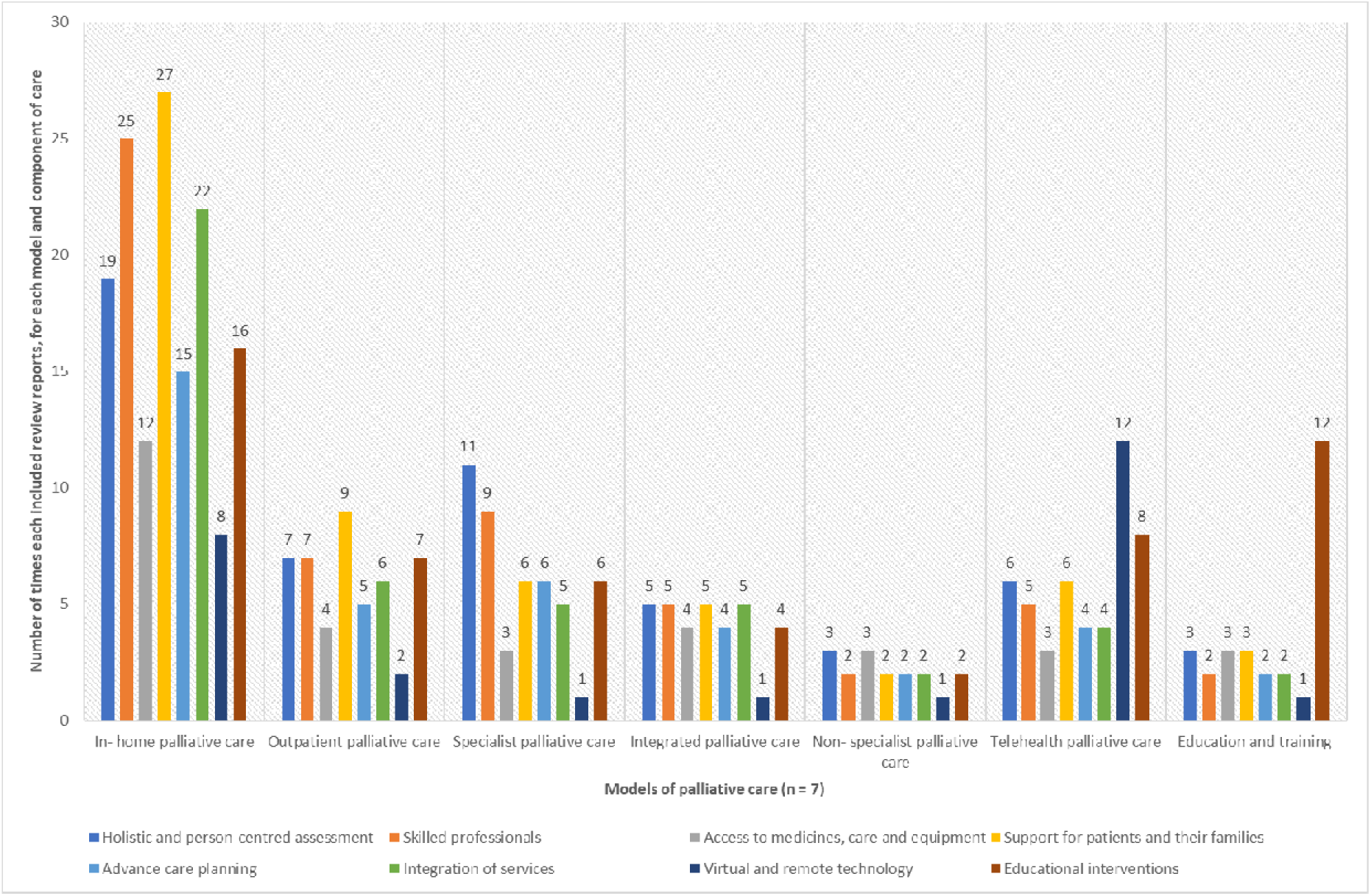
Identified components of care within the included reviews and their frequency across the identified models of palliative care **Footnote**: Included reviews which report on more than one model and/or component are counted more than once

### Outcomes of care

Twenty-four outcomes of care (see ***Table 4***) were identified from all included reviews (these were reported by n = 55; 83.3% of included reviews); we categorised these into four groups:

a. **patient outcomes** (reported by 46 reviews across all models of care; 69.7% of included reviews): symptoms (general), behavioural symptoms, psychological distress, functional status, quality of life, advance care planning, survival, satisfaction with care, coping, adherence and autonomy, and patient unmet needs;
b. **family or informal caregiver outcomes** (reported by 21 reviews across all models of care; 31.8% of included reviews): include caregiver behavioural symptoms, caregiver psychological distress, caregiver grief, caregiver quality of life, caregiver burden, caregiver self-efficacy/competence, caregiver satisfaction with care, and caregiver unmet needs;
c. **professional outcomes** (reported by 10 reviews across all models of care; 15.2% of included reviews): professional competence, and professional’s satisfaction with care;
d. **service utilisation and costs** (reported by 34 reviews across all models of care; 51.5% of included reviews): place of death, hospitalisation, emergency department visits, and overall or specific healthcare costs.

**Table 4:** Summary of reported evidence from the included reviews, grouped by model of care.

| Models of palliative care | Outcome categories | Specific outcomes | Narrative synthesis of evidence |
| --- | --- | --- | --- |
| **Model 1** (In-home care) | Patient outcomes | Symptoms (general) | **Evidence of positive effect:** Pain and symptom management<sup>11, 13, 50, 65, 90</sup> including breathlessness<sup>78</sup> were improved for patients receiving in-home care services, especially with multi-professional<sup>45, 76, 99</sup> and specialist palliative care teams providing the care.<sup>24*</sup><br>**Evidence of negative effect:** Two reviews found that in-patient care regulated symptoms better than home based care.<sup>13, 87</sup><br>**Evidence of no effect:** For chronic respiratory patients, there was no evidence of symptom improvement.<sup>78</sup> |
|  |  | Behavioural symptoms | **Evidence of positive effect:** Compared to usual care, home palliative care was more effective in reducing behavioural disturbances.<sup>68</sup> |
|  |  | Functional status | **Evidence of positive effect:** Functional status was improved for patients receiving in-home care in comparison to usual care.<sup>11, 76</sup> |
|  |  | Quality of life | **Evidence of positive effect:** Quality of life including social quality of life in patients receiving in-home care improved in comparison to usual care.<sup>11, 40, 45, 50, 66, 76, 82, 90</sup><br>**Evidence of no effect:** Two reviews showed that palliative home care teams and specialist nurses had little or no effect on quality of life for patients dying at home with no consistent difference found between specialist and usual care.<sup>15, 87</sup> In chronic respiratory patients, no significant benefit was found.<sup>78</sup> Similarly, GP-led advance care planning with social care support showed no quality of life benefits.<sup>79*</sup> |
|  |  | Advance care planning | **Evidence of positive effect:** End-of-life patients and their caregivers preferred home visits to better achieve both medical and non-medical preferences,<sup>43</sup> and achieved increased numbers of goals of care discussions and documentations of advance care planning, compared to usual care,<sup>45, 78</sup> with improved staff-older person relationships<sup>73*</sup>. |
|  |  | Satisfaction with care | **Evidence of positive effect:** Satisfaction improved for patients receiving in-home care in comparison to usual care,<sup>11, 45, 58, 59, 80, 83, 91</sup> particularly where care was delivered by multi-professional<sup>99</sup> or specialist teams,<sup>45</sup> which foster familiarity, and improved patient-professional relationships.<sup>96</sup><br>**Evidence of negative effect:** Absence of regular communication with doctors, nurses including primary and secondary care staff resulted in uncertainty as to who should be contacted in times of needs thus negatively affecting satisfaction with care.<sup>93</sup> |
|  |  | Coping, adherence, and autonomy | **Evidence of positive effect:** Coping and self-efficacy were improved for patients receiving home-based interventions compared to usual care.<sup>40, 58</sup> |
|  |  | Patient unmet needs | **Evidence of positive effect (in identifying unmet needs):** Unmet needs such as communication, spiritual, psychosocial, practical, informational and respite needs as well as those associated with isolation and loss of autonomy were better identified.<sup>93</sup> Lack of identification of these unmet needs negatively impacted the care delivered to patients and carers.<sup>93</sup> |
|  | Family or informal caregiver outcomes | Caregiver psychological distress | **Evidence of positive effect:** Caregivers who received bereavement support showed decreased depressive symptoms and increased resilience.<sup>70</sup> |
|  |  | Caregiver grief | **Evidence of no effect:** One review found no effect on caregiver grief for caregivers who looked after patients receiving home-based palliative care in comparison to usual care.<sup>13*</sup> |
|  |  | Caregiver quality of life | **Evidence of positive effect:** Increased quality of life/ caregiver outcomes were associated with in-home care compared to usual care.<sup>11, 59</sup> |
|  |  | Caregiver burden | **Evidence of positive effect:** Home visits provided respite for caregiver and reduced burden.<sup>92</sup><br>**Inconsistent evidence:** The evidence is mixed about the effect of in-home care on caregiver burden compared to usual care, with some studies finding little or no effect.<sup>13*</sup> |
|  |  | Caregiver satisfaction with care | **Evidence of positive effect:** Home care improved caregiver satisfaction with care compared to usual care<sup>11, 58, 77</sup> through the provision of information or education to families and patients receiving the care.<sup>70, 83*</sup><br>**Inconsistent evidence:** There is mixed evidence about the effects of home-based end-of-life care on caregiver satisfaction with care as this was found to disappear after six months.<sup>80*</sup> |
|  |  | Caregiver unmet needs | **Evidence of negative effect:** Where unmet caregiver needs were not identified, there was increase in feelings of isolation, and some loss of activities (e.g. attend religious services, travel, etc.) attributed to home-based palliative care.<sup>93</sup> |
|  | Professional outcomes | Professionals satisfaction with care | **Evidence of positive effect:** Positive experiences were reported by medical professionals for community-based palliative care.<sup>58</sup><br>**Inconsistent evidence:** Physicians reported it was difficult to decide on whether and when to hospitalise patients' appropriately whilst providing care at home for the patients.<sup>43</sup> |
|  | Service utilisation and costs | Place of death | **Evidence of positive effect:** Home was associated with a more peaceful death.<sup>74</sup> Patients who accessed out-of-hours services were more likely to die at home than non-users.<sup>11, 24*</sup> Access to home nursing increased the likelihood of dying at home<sup>13, 43, 45, 50, 53, 58, 65, 66, 68, 80, 82, 97</sup> and this did not compromise quality of life or cost.<sup>10</sup> |
|  |  | Hospitalisation | **Evidence of reduced service use:** Provision of in-home respite care reduced hospitalisations<sup>66</sup> including length of hospitalisation.<sup>43, 45, 50, 53, 58, 77, 82</sup> Availability of, and access to out-of-hours services for patient's family and caregivers enabled continued care at home, and increased comfort/less isolation.<sup>54, 60, 91</sup><br>**Inconsistent evidence:** There was substantial variability in the reported impact of in-home care on admission to hospital for home-based palliative care.<sup>80*</sup><br>**Evidence of no effect:** No significant evidence for impact of palliative care on use of health services for chronic respiratory patients.<sup>78</sup> |
|  |  | Emergency department (ED) visits | **Evidence of reduced ED visits:** Home-based palliative care is associated with fewer emergency department visits,<sup>50, 58, 82, 91</sup> and major reduction in inpatient admissions compared to usual care.<sup>84</sup><br>**Evidence of no effect:** One review reported that ED usage was equivalent between those with access to 24-hour specialist palliative care and without.<sup>24*</sup> |
|  |  | Overall or specific healthcare costs | **Evidence of reduced costs:** There was a reduction in healthcare use or cost for patients accessing in-home care compared to cost before access, or usual care<sup>11, 53, 59, 65, 77, 81, 82, 84, 91, 99</sup> with hospital costs being lower for this<br>cohort.<sup>24*</sup> Provision of in-home respite care, physician-led home-based care reduced hospital admission costs.<sup>66</sup><br>**Inconsistent evidence:** There is mixed evidence around cost-effectiveness of home-based palliative care. For example, Shepperd et al reported that one trial reported no differences in overall net health costs while a second trial reported that the mean cost per day incurred by those participants receiving home-based care was lower than for those receiving standard care.<sup>13, 80*</sup><br>**Evidence of increased costs:** Home care services were approximately 30% more costly during the last 24 months of illness than hospice or conventional care.<sup>97</sup> |
| **Model 2**<br>(Outpatient care) | Patient outcomes | Symptoms (general) | **Evidence of positive effect:** Outpatient care provided in the home or clinic by palliative care specialists alleviated patient's symptoms.<sup>10, 46, 91</sup> |
|  |  | Psychological distress | **Evidence of positive effect:** There were reported improvements to patient wellbeing for patients receiving outpatient care in the home or community settings.<sup>91</sup> |
|  |  | Quality of life | **Evidence of positive effect:** Outpatient care provided in the home or clinic by palliative care specialists improved quality of life.<sup>46, 91</sup><br>**Inconsistent evidence:** There were insufficient studies with conclusive results on patient health-related quality of life for outpatient care provided by palliative care specialists (mostly nurse-led).<sup>98</sup> |
|  |  | Survival | **Evidence of no adverse effect on survival:** There was no significant difference between outpatient care and usual care for patient survival.<sup>91</sup> |
|  |  | Satisfaction with care | **Evidence of positive effect:** Outpatient care provided in-home or community settings improved patient's satisfaction.<sup>91</sup> |
|  | Family or informal caregiver outcomes | Caregiver psychological distress | **Evidence of positive effect:** There was decreased psychological distress for caregivers following outpatient care interventions. However, upon follow-up at 4 and 8 months and after bereavement, this benefit did not persist.<sup>40, 87</sup> |
|  |  | Caregiver quality of life | **Evidence of positive effect:** Caregiver's physical, emotional, or social quality of life domains were improved by outpatient care interventions.<sup>40</sup> |
|  |  | Caregiver burden | **Evidence of positive effect:** Outpatient care delivered reductions in objective caregiver burden (specifically caregiver responsibility and perceived disturbances in aspects of their life).<sup>40, 46, 91</sup> |
|  |  | Caregiver self-efficacy/competence | **Evidence of positive effect:** Outpatient care interventions delivered in-home improved caregiver's self-efficacy/competence, although this effect was not sustained (at 2 and 6 months).<sup>40</sup> |
|  |  | Caregiver satisfaction with care | **Evidence of positive effect:** Caregivers and family members of patients receiving outpatient care in their home reported increased satisfaction after 1 month of receiving care.<sup>91</sup> |
|  |  | Caregiver unmet needs | **Evidence of positive effect:** There were fewer unmet needs reported by caregivers and family members of patients receiving outpatient care in the home or community settings.<sup>91</sup> |
|  | Professional outcomes | Professionals satisfaction with care | **Evidence of positive effect:** Outpatient care was associated with improved physician satisfaction, partly because of time savings.<sup>91</sup> |
|  | Service utilisation and costs | Place of death | **Evidence of positive effect:** Provision of outpatient care in-home or community settings improved the likelihood of dying at home where this was preferred.<sup>24, 50, 52, 66</sup> |
|  |  |  | **Inconsistent evidence:** Two reviews reported inconclusive results about place of death.<sup>10, 91</sup> |
|  |  | Hospitalisation | **Evidence of reduced use:** Outpatient care provided in the home or clinic by palliative care specialists reduced rehospitalisation compared to usual care.<sup>46, 91</sup> |
|  |  | Emergency department visits | **Evidence of reduced visits:** Fewer emergency department visits were reported where patients received outpatient care in the home or community settings.<sup>91</sup> |
|  |  | Overall or specific healthcare costs | **Evidence of reduced costs:** One review indicated that outpatient care provided in-home reduced cost of services depending on the interventions applied,<sup>91</sup> and there were positive benefits without compromising costs.<sup>10</sup><br>**Inconsistent evidence:** Varied results from some studies, with reduction in cost of care from some, but no difference from others, in comparison to usual care.<sup>87</sup> |
| **Model 3**<br>(Specialist care) | Patient outcomes | Symptoms (general) | **Evidence of positive effect:** Home-based specialist palliative care improved symptoms.<sup>11, 15, 45, 46, 70, 90, 94</sup><br>**Inconsistent evidence:** There were mixed results among studies assessing control of patient symptoms.<sup>71, 88</sup> |
|  |  | Psychological distress | **Evidence of positive effect:** Specialist palliative care (SPC) intervention delivery via telehealth improved psychological distress including reductions in anxiety and depression.<sup>88</sup> SPC interventions yielded moderate benefits in emotional wellbeing, especially after receiving care over at least 3 months.<sup>61*</sup> |
|  |  | Functional status | **Evidence of positive effect:** Functional status was improved for patients with in-home support from caregivers who had access to support from SPC professionals, compared to usual care.<sup>11</sup> |
|  |  | Quality of life | **Evidence of positive effect:** SPC interventions yielded substantial benefits in quality of life for receiving patients from 2 weeks,<sup>61</sup> and improved overall, the quality of life for patients who received home-based SPC compared to those who did not.<sup>11, 15, 45-47, 90</sup><br>**Inconsistent evidence:** When comparing effectiveness of telehealth interventions with SPC providers, compared to standard care, the results were inconsistent.<sup>88</sup> |
|  |  | Satisfaction with care | **Evidence of positive effect:** SPC services improved patient satisfaction<sup>10</sup> especially for those with access to telehealth support within 24 hours<sup>16, 55, 56, 89, 95</sup> including those in the community.<sup>82</sup> SPC interventions also addressed multiple needs and coordination was key.<sup>48*</sup><br>**Inconsistent evidence:** There were mixed results in studies reporting satisfaction with care after delivery of SPC led telehealth intervention.<sup>88</sup> |
|  |  | Coping, adherence, and autonomy | **Evidence of positive effect:** SPC-led interventions increased knowledge and skills of patients and relatives compare to before the intervention was provided.<sup>45, 83*</sup> These interventions also led to better treatment adherence from patients compared to usual care.<sup>45, 76</sup> |
|  | Family or informal caregiver outcomes | Caregiver quality of life | **Evidence of positive effect:** SPC led interventions improved caregivers' quality of life.<sup>11</sup> |
|  |  | Caregiver satisfaction with care | **Evidence of positive effect:** SPC led interventions delivered via telehealth,<sup>94</sup> or provided in-home increased satisfaction for caregivers.<sup>11, 83*</sup> |
|  |  | Caregiver unmet needs | **Evidence of positive effect:** There were fewer unmet needs reported by caregivers and family members of patients receiving SPC care in the home or community settings.<sup>91</sup> |
|  | Professional | Professionals | **Evidence of positive effect:** Collaboration and shared learning among SPC teams increased productivity and |
|  | outcomes | satisfaction with care | enhanced care delivery.<sup>45</sup> |
|  | Service utilisation and costs | Place of death | **Evidence of positive effect:** Access to home-based SPC increased the proportion of people who died at home,<sup>15, 43, 53, 65, 68, 80*</sup> and increased chances of death in patient's preferred location.<sup>11, 70</sup> |
|  |  | Hospitalisation | **Evidence of reduced use:** SPC teams, with their expertise, and coordinated approach, helped reduce hospital admissions, hospitalisation, and length of stays,<sup>10, 45, 46, 53, 66</sup> especially when SPC telehealth interventions were fully implemented.<sup>88</sup> |
|  |  | Overall or specific healthcare costs | **Evidence of reduced costs:** Access to coordinated SPC reduced total costs and overall healthcare use for patients<sup>11, 15, 69, 84</sup> and can lead to cost savings for long term home-based care patients.<sup>66</sup><br>**Inconsistent evidence:** There were mixed effects of SPC interventions on cost savings in some reviews.<sup>10, 65, 80*</sup> |
| **Model 4**<br>(Integrated care) | Patient outcomes | Symptoms (general) | **Evidence of positive effect:** Integrated care reduced the symptom burden of palliative care patients; however, the details of how integration was achieved were not always clear.<sup>99</sup> Patients receiving combined primary care (general practitioner) and secondary care interventions had improved pain management and symptom control.<sup>86</sup><br>**Evidence of no effect:** Patients receiving integrated care showed no improvement in symptom burden after 1-3 months,<sup>52</sup> even when a videoconference platform was used for care delivery<sup>55</sup> and care was provided in the community by multidisciplinary teams.<sup>82</sup> |
|  |  | Psychological distress | **Evidence of positive effect:** Integrated palliative care lessened one or more depression symptoms.<sup>52, 55</sup> |
|  |  | Functional status | **Evidence of positive effect:** Integrated care delivered by specialist physician in-home through telehealth appointments reduced functional decline.<sup>66</sup> The functional states of patients receiving integrated care were better maintained.<sup>86</sup> |
|  |  | Quality of life | **Evidence of positive effect:** Integrated palliative care interventions improved quality of life<sup>55, 82</sup> with patients with lung cancer benefitting the most compared to those with gastrointestinal cancer.<sup>52</sup><br>**Evidence of negative effect:** At 6–12 months, quality of life was not improved by integrated palliative care interventions.<sup>52</sup> |
|  |  | Advance care planning | **Inconsistent evidence:** The results were mixed on the benefits of integrated care on advance care planning for community-based patients<sup>82</sup> even with advance care plan directives in place.<sup>43</sup> |
|  |  | Survival | **Inconsistent evidence:** There was no clear evidence of whether integrated palliative care had any association (positive or negative) with survival.<sup>52, 78</sup><br>**Evidence of no effect:** There was no difference in time to death between integrated home-based care and hospital-based care.<sup>86</sup> |
|  | Family or informal caregiver outcomes | Caregiver quality of life | **Inconsistent evidence:** Studies that assessed caregiver quality of life in relation to community-based palliative care were limited and the evidence varied.<sup>82</sup> |
|  |  | Caregiver satisfaction with care | **Evidence of positive effect:** Access to home-based palliative care improved caregiver satisfaction by providing support, educational resources, information, and supporting patient preferences.<sup>43, 87</sup> |
|  | Service utilisation and costs | Place of death | **Evidence of positive effect:** Studies reported increase in the proportion of patients who died at home and a reduction in hospital deaths when they had access to integrated home-based palliative care programmes delivered by specialists.<sup>43, 53, 80, 82</sup> |
|  |  | Hospitalisation | **Evidence of reduced use:** Hospital admissions and length of hospital stay were reduced by integration of primary and secondary services in the provision of in-home palliative care.<sup>53, 86</sup> Integration of virtual consultations and provision of in-home respite care and physician-led home-based palliative care helped achieve this as well.<sup>66</sup><br>**Inconsistent evidence:** Service utilisation was reported inconsistently across studies, including emergency department and hospitalisation use.<sup>52</sup> |
|  |  | Emergency department visits | **Evidence of reduced visits:** Patients who received home-based palliative care had significantly less emergency department admissions compared to usual care.<sup>53, 86</sup> |
|  |  | Overall or specific healthcare costs | **Evidence of reduced costs:** Cost reduction was most noticeable closer to death when home-based palliative care was used compared to standard care.<sup>53, 70, 99</sup> Integrated care interventions was associated with a lower mean total cost per day.<sup>52</sup><br>**Inconsistent evidence:** When healthcare costs were assessed per country, studies found inconsistent results.<sup>82</sup> |
| **Model 5 (Non-specialist care)** | Patient outcomes | Symptoms (general) | **Evidence of no effect:** Non-specialist palliative care delivered in a community setting did not reduce symptom frequency or lessen symptom severity.<sup>70</sup> |
|  | Family or informal caregiver outcomes | Caregiver psychological distress | **Evidence of positive effect:** Non-specialist psychosocial or educational interventions provided to caregivers decreased caregiver psychological distress including anxiety.<sup>40, 94</sup> |
|  |  | Caregiver quality of life | **Evidence of positive effect:** Overall quality of life improved for caregivers, when non-specialist psychosocial or educational interventions were provided.<sup>40</sup><br>**Evidence of no effect:** There was no significant difference between telehealth interventions provided by non-specialist healthcare providers for caregiver's quality of life.<sup>94</sup> |
|  |  | Caregiver burden | **Evidence of positive effect:** Caregivers experienced lessened disruptions in their daily lives and reduced objective burden when non-specialist psychosocial or educational interventions were provided.<sup>40</sup><br>**Inconsistent evidence:** There were varied results from studies about caregiver burden with some showing decreased burden and some not, when telehealth interventions were provided.<sup>94</sup> |
|  |  | Caregiver self-efficacy/competence | **Evidence of positive effect:** Self-efficacy for caregivers improved when non-specialist psychosocial or educational interventions were provided but this was sustained.<sup>40</sup> |
|  | Professional outcomes | Professionals satisfaction with care | **Evidence of positive effect:** Telehealth education interventions increased the confidence levels of the healthcare providers.<sup>95</sup><br>**Evidence of negative effect:** As a result of limited palliative care knowledge, home healthcare physicians struggled with hospitalisation decisions and were less likely to initiate community-based palliative care.<sup>43</sup> Generalists such as district nurses identify particular issues because of the difficulties of providing palliative care amongst all the other calls on a district nurses time.<sup>96</sup> |
|  | Service utilisation and costs | Place of death | **Evidence of positive effect:** Patients enrolled in a non-specialist community-based palliative care intervention were more likely to die in their location of choice and less likely to die in hospital than those not enrolled.<sup>70</sup> |
|  |  | Hospitalisation | **Evidence of reduced use:** Home-based care delivered by nurse practitioners demonstrated substantial reductions in emergency department visits, hospitalisations, long-term-care admissions and bed days.<sup>84</sup> |
|  |  | Overall or specific healthcare costs | **Evidence of reduced costs:** Home-based care delivered primarily by nurse practitioners reduced hospital costs.<sup>84</sup> |
| **Model 6**<br>(Telehealth care) | Patient outcomes | Symptoms (general) | **Evidence of positive effect:** Provision of telecommunication or virtual technology interventions improved physical symptoms, symptom indicators, symptom distress and burden compared to usual care. <sup>16, 44, 47, 55, 88</sup><br>**Evidence of no effect:** One review reported that provision of telecommunication or virtual technology interventions did not improve symptom distress. <sup>44</sup> |
|  |  | Psychological distress | **Evidence of positive effect:** Telecommunication interventions were more effective at identifying distress as opposed to regular psychiatric assessments. <sup>16, 95</sup> |
|  |  | Functional status | **Evidence of positive effect:** Physical, mental and social function were improved (statistically significant) for those using telecommunication interventions compared to usual care. <sup>44</sup><br>**Evidence of no effect:** Compared to usual care, there was no significant between-group effects for functional status at 12 weeks following home-based palliative heart failure intervention. <sup>44</sup> |
|  |  | Quality of life | **Evidence of positive effect:** Telehealth palliative care interventions, specifically telephone consultations, showed overall increased quality of life compared to usual care. Specific areas of improvement identified were health, physical, mental and social related quality of life. <sup>44, 47, 82</sup> |
|  |  | Advance care planning | **Evidence of no effect:** While patient preferences, uncertainty, and comfort care choices were reflected in advance care planning indicators, there was no positive effect of telehealth educational videos on advance care planning. <sup>44</sup> |
|  |  | Satisfaction with care | **Evidence of positive effect:** Enhanced access to care and convenience, in the context of telepalliative care and out-of-hours telephone support, are highly valued by patients and families. <sup>57, 58, 60</sup> |
|  | Family or informal caregiver outcomes | Caregiver quality of life | **Evidence of positive effect:** Video consultations showed improvements in caregivers' quality of life. <sup>47</sup> |
|  |  | Caregiver satisfaction with care | **Evidence of positive effect:** Out-of-hours phone support reduced isolation, offered reassurance and provided a sense of security for caregivers. <sup>55, 60</sup> |
|  | Service utilisation and costs | Overall or specific healthcare costs | **No evidence:** There was little or no evidence on the costs or cost effectiveness of telephone advice lines. <sup>60</sup> |
| **Model 7**<br>(Education and training for delivering care) | Patient outcomes | Symptoms (general) | **Evidence of positive effect:** Symptom control improved when home care services and psychosocial interventions were delivered by multi-professional teams trained in palliative care. <sup>68, 99</sup> |
|  |  | Survival | **Evidence of positive effect:** Nurse-led education improved survival and telemonitoring was associated with reduced all-cause mortality compared to usual care. <sup>56, 66</sup> |
|  |  | Satisfaction with care | **Evidence of positive effect:** Compared to usual care, in-home care with multi-professional teams trained in palliative care increased satisfaction with care and positive feedback. <sup>99</sup><br>**Evidence of negative effect:** Patients expressed a want for information regarding their illnesses and this led to dissatisfaction with care. <sup>93</sup> Nurse-led education did not improve satisfaction with care. <sup>66</sup> |
|  | Family or informal caregiver outcomes | Caregiver behavioural symptoms | **Evidence of positive effect:** One study found that primary caregivers of Alzheimer's patients who took a palliative care training course had long-term positive effects on managing the patients' behavioural symptoms, with improvements observed at both 12 and 28 months. <sup>68</sup> |
|  |  | Caregiver | **Evidence of positive effect:** Caregiver psychosocial support programs and educational interventions decreased |
|  |  | psychological distress | caregiver psychological distress.<sup>40, 82</sup> |
|  |  | Caregiver quality of life | **Evidence of positive effect:** The physical and social domains of quality of life and emotional aspects of quality of life were significantly improved for caregivers following educational interventions.<sup>40</sup> |
|  |  | Caregiver burden | **Evidence of positive effect:** Caregiver burden was reduced for caregivers involved in education/training interventions.<sup>40</sup> |
|  |  | Caregiver self-efficacy/competence | **Evidence of positive effect:** Caregiver self-efficacy/competence was improved by educational interventions and information about out-of-hours telephone advice lines as it increased their preparedness.<sup>40, 58, 60, 83*</sup> |
|  |  | Caregiver satisfaction with care | **Evidence of positive effect:** Provision of information about disease progression, behaviours, emotions and physical symptoms to expect, all improved caregiver satisfaction with care.<sup>70, 80, 83*</sup> |
|  | Professional outcomes | Healthcare professional competence | **Evidence of positive effect:** Staff education and informational resources for palliative care staff facilitated by videoconferencing, improved knowledge scores and self-efficacy of professionals, and promoted improved patient-nursing staff relationships.<sup>56, 70, 73, 82</sup> Conversely, district nurses felt unprepared to address psychological issues due to a lack of training.<sup>96</sup> |
|  | Service utilisation and costs | Hospitalisation | **Inconsistent evidence:** There were mixed results in terms of number of admissions and time to first hospitalisation for patients who accessed telemonitoring; however, this cohort were generally older in comparison to usual care, which may account for some of the inconsistencies in the evidence.<sup>56</sup> |

*\*: Indicates high-quality reviews*

The majority of the patient outcomes were reported in relation to the in-home (26 reviews), and specialist (21 reviews) models of care. The family or informal caregiver outcomes were mostly reported in the in-home (10 reviews) and education and training (8 reviews) models of care (where education or training was provided to families). Professional outcomes were mostly reported in the education and training (5 reviews) model (where education and training were provided to professionals). Service utilisation and cost outcomes were mostly reported in relation to the in-home (25 reviews) model of care. Across all included reviews and models of care, the most common outcomes (reported by ≥ 20 reviews, and ordered by most common first) were: symptoms (general); satisfaction with care; overall or specific healthcare costs; quality of life; place of death; and hospitalisation.

Positive benefits were most commonly reported in relation to patient outcomes for in-home and specialist models of care; relatively less evidence was reported for the non-specialist model of care. For family or informal caregiver and professional outcomes, these were most commonly reported in relation to the education and training model of care, where education or training was delivered to these groups. Evidence of changes in service utilisation or costs were reported for the in-home and specialist models of care, but less often that for patient outcomes. See ***Figure 3*** and ***Table 4*** for further details of the direction and volume of evidence.

**Figure 3:**
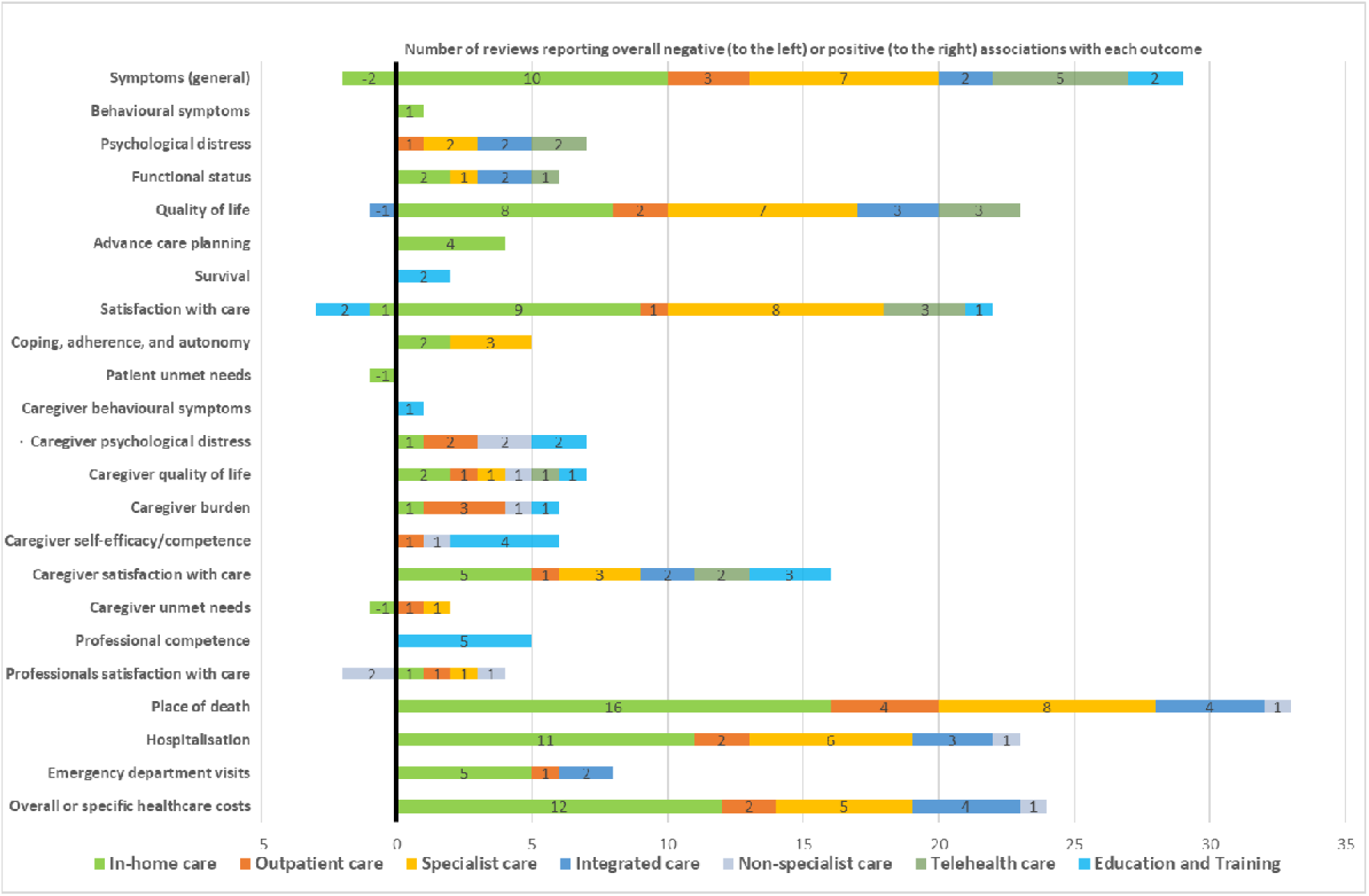
Illustration of the identified benefits and associations between some of the models of palliative care and outcomes from the included reviews **Footnote:** Included reviews which report on more than one outcome are counted more than once

All the identified models of care had some reported evidence of a positive effect, mostly in relation to patient outcomes, and service utilisation and costs. **Figure 3** provides an overview of the numbers of reviews reporting each outcome, by model of care, and the reported direction of effect. Caution should be exercised in interpreting figure 3, given that inconsistent findings are not captured in the figure, there may be some ‘amplification’ where individual studies are reported by more than one review, and the quality appraisals of reviews are not incorporated. However, it does demonstrate where most research has been conducted, and gives a headline indication of the direction of effects identified.

In general, we found evidence on service utilisation, costs and cost-effectiveness very limited. Several studies examined the overall healthcare costs of various interventions in the different models of care using outcomes such as overall costs, specific hospital costs, cost effectiveness, hospital utilisation, and impact of place of death on costs. For example, an increase in the likelihood of dying at home was shown for the in-home model of care which included access to home nursing compared to alternative models ^13, 43, 45, 50, 53, 58, 65, 66, 68, 80, 82, 97^ and this did not compromise quality of life ^10^. Home-based specialist palliative care from specialist nurses increased home deaths without compromising symptoms. ^15, 65^ Comprehensive and robust evidence on the cost implications of telephone advice lines for telehealth models of care related to the benefits is ‘very limited’, but there is some limited indication that costs can be reduced through out-of-hours telephone advice lines ^60^ and videoconference-based palliative care ^67^. Janke et al ^59^, in a non-cancer population where most were home-based, suggests that palliative care is ‘cost-saving or at least cost-neutral’. Spencer and colleagues ^81^ in their review conducted from the perspective of the healthcare system, showed that hospital-based palliative care costs are higher than hospice or home-based palliative care (however, patient and informal caregiver costs were not included).

## Discussion

### Main findings of the review

In this extensive and complex systematic review of review-level evidence about palliative and end-of-life care for people at home, we have been able to draw together a large volume of evidence and present an overarching picture of the evidence for models of palliative and end-of-life care for those at home.

First, we provide insights into the existing models and components of in-home palliative care. We identified seven models, distinguished by setting (where care is delivered; either at home or through outpatient care), who is delivering the care (type of professionals; either specialist, integrated with specialist, or non-specialist palliative care), and mode of delivery (telehealth or education/training). Not all models are equivalent: some only represent part of care e.g. outpatients, telehealth, education/training. It is clear that two models - the outpatient model, and the model delivered by non-specialists in palliative care - have been much less researched.

Second, we synthesise the overall evidence, and show that this largely supports in-home palliative care, especially if delivered via specialist palliative care models or integrated palliative care models (integration is between specialist and non-specialist services). Careful interpretation of our findings (see **Table 4**) shows surprising consistency in the evidence; consistency between included reviews (as evidenced by the same direction of effect for a number of reviews for the same outcomes) and consistency in the outcomes showing benefit (for instance, the positive effect of several models on patients’ symptoms, psychological distress, and functional status, in contrast to the more mixed evidence across models on patients’ quality of life, where it is perhaps harder to show impact). This consistency is also apparent across the higher-quality reviews. The in-home and specialist models of care have most evidence for reducing patients’ symptoms, overall or specific healthcare costs, attaining patient, family, and professional satisfaction with care, and reducing hospitalisation. This is supported by a recent meta-analysis and meta-regression on specialist palliative care.^61^ There is less evidence for non-specialist palliative care, but there may also be more variation in this model, accounting for less consistency in what evidence there is (see **Table 4**). In addition, the education and training model extensively supports family or informal caregivers in delivery of care, with evidence suggesting training, increases their competence, quality of life, and reduces psychological distress; an important finding given that 1 in 10 carers report unwillingness to care again.^100^

The in-home care model sometimes showed inconsistent results for service utilisation and cost (see **Table 4**), such as emergency department visits, hospitalisation, and overall healthcare costs. One review indicated that care at home may be approximately 30% more costly in the last 24 months of illness than hospice or conventional care. ^97^ and it is likely that this reflects different cost horizon and perspectives. Despite this, achieving preferred place of care and death, improved overall healthcare costs, symptom relief, quality of life and wellbeing, alongside satisfaction, by the identified models of care, especially the in-home, and specialist models of care, is in line with recent findings, ^50, 54, 57–59, 61, 66, 73, 74, 79^ and may reflect affordability, and meeting preferences of patients and family to care for and die in a place of comfort – home.

Considering the benefits that was observed for patients, ^16, 44, 47, 53, 57, 58, 60, 82, 88, 95^ and caregiver ^47, 55, 60^ outcomes, supporting professionals in effective service delivery, and provision of care services to support individuals to be cared for – in their own homes – through telehealth care is promising. However, there is limited evidence to support effectiveness of specific services such as telephone advice lines, and out-of-hours telephone lines. ^60^ There has been little study of the effects of telephone support and telehealth in general on symptom management, functional status, and advance care planning. ^44^ This requires more investigation as it may support wellbeing while reducing overall costs.

### Strengths and limitations of the review

We have endeavoured to present an overview of a large volume of evidence, and at the same time provide sufficient detail to identify the sources of evidence for any one model of care and related outcomes. We used two previous meta-level reviews ^10, 25^ on models, components and outcomes of palliative care to inform our analysis. We used narrative synthesis to address the heterogeneity that results from including multiple types of reviews without a uniform model of care or standard outcomes. There is a challenging trade-off between synthesis and detail. Strengths of this review include: registration of, and adherence to a study protocol including reporting changes; detailed search strategy developed by specialist; dual reviewer screening; quality appraisal; and a focus on more recent evidence given most included reviews were less than 10 years old. Limitations include: restriction to English-language searches; no exclusion of low-quality review articles (although we report high quality reviews in **Table 4**); potential publication bias (with positive findings more likely to be published). Our application of narrative synthesis exposed our analysis to inherent subjectivity that can affect the reproducibility and validity of our findings. However, we applied a critical reflexive approach in all stages of the research, were transparent in our reporting, and adopted systematic techniques and PRISMA guidance to control for this bias. We found it was not possible to explore directly, the associations between components of care and outcomes as earlier planned. It is worthy of note that most of the included reviews were from high-income countries and findings may not be generalisable to low or middle-income countries. There is also a moderate risk of an ‘amplification’ effect on some evidence, as some reviews may report the same individual studies and we could not report overlap between reviews at the individual study level.

### What this review adds and implications for practice, policy and research

This review of reviews provides an overview of a large volume of evidence with sufficient detail to understand current evidence on any one model of care, components, and related outcomes. The evidence most strongly supports the provision of in-home palliative care, specialist palliative care, and integrated services, with positive effects largely reported on patient outcomes for these models of care.

Future experimental and observational studies are required to assess the cost effectiveness of the identified models, components, and outcomes of care in different economic regions and healthcare systems. System wide research is particularly required. Also, research is needed to standardise outcomes of care and explore multidisciplinary and integrated approaches to provision of holistic care.

## Conclusion

This meta-level evidence most strongly supports the provision of in-home palliative care, as this is where there is most review level evidence showing positive effect on patient outcomes. There was also evidence to support specialist palliative care and integrated teams (integration of primary palliative care with specialist support), and education and training for both informal family carers and professionals.

## Differences between protocol and review

We made minor revisions to the title and objectives from the protocol to streamline the focus of this review, along with refining the inclusion and exclusion criteria, outcomes, and search strategy to improve findings. We made several minor changes to the style and organisation of the text to improve the clarity and conciseness.

Also, the extracted information and approach to quality appraisal were updated to include additional elements aimed at improving the quality of the review.

We mapped the models to outcomes, not components as previously planned but endeavoured to reflect on how the models and components relate to the outcomes.

## Supporting information

Supplemental material 3: Data extraction template.

Supplemental material 1: Full search strategies for original main database searches.

Supplemental material 2: Characteristics of included reviews.

## Data Availability

All data produced in the present work are contained in the manuscript

## Author’s contributions

SP, FEMM, KES, SB and IJH were responsible for conceptualisation and review design, with critical input from AM, RLC, SG, TJ and AEB. SG and SP developed and refined the search strategy. SP, CO, AM, RP, JY, TC, SH, IW, AS, RLC and SG were responsible for review identification, screening, inclusion and extraction of data. SP, CO, RP, JY, TC, SH, IW, AM, TJ, AEB, SB, IJH, KES and FEMM were involved in the analysis and interpretation of results. SP, CO, RP and FEMM were involved in preparing the draft manuscript. All authors reviewed the results and approved the final version of the manuscript.

## Acknowledgements and funding

This study was conducted as part of the Better End of Life Programme, which is funded by Marie Curie (grant MCSON-20-102) and awarded to KES, IJH, FEMM and SB. The research was carried out by King’s College London in collaboration with Hull York Medical School, University of Hull, and University of Cambridge. The funder was not involved in the study design, data collection and analysis, or interpretation of results. KES is the Laing Galazka Chair in palliative care at King’s College London, funded by an endowment from Cicely Saunders International and the Kirby Laing Foundation. IJH is an NIHR Senior Investigator Emeritus. FEMM is a National Institute for Health Research (NIHR) Senior Investigator. IJH and SB are supported by the NIHR Applied Research Collaboration (ARC) South London (SL) and NIHR ARC East of England, respectively. SG’s time was funded by Yorkshire Cancer Research under the TRANSFORM programme (award reference HEND405SPT). The views expressed in the report are those of the authors and not necessarily those of the NIHR, or the Department of Health and Social Care.

## Research ethics and patient consent

This study did not require ethics approval as it was a review of already published reviews.

