## Supplemental material 3: Data extraction template. for "Models, components, and outcomes of palliative and end-of-life care provided to adults living at home: A systematic review of reviews"

| **Field** | **Extracted information** |  | **Example and guidance on how to complete** | **Additional notes, comments or actions required** |
| --- | --- | --- | --- | --- |
| Person extracting the data (Initials) |  | | *First name and surname initials (e.g. SP)* |  |
| Date form was completed |  | | *e.g. 01/12/2021* |  |
| **Publication details and review characteristics** | | | | |
| Review title |  | | *Include the full title of the review* |  |
| Author(s) |  | | *Include all review authors. Example format: Bainbridge D, Hsien S & Sussman J* |  |
| Year |  | | *e.g. 2016* |  |
| Study ID |  | | *e.g. Bainbridge et al. (2016) J Am Geriatr Soc* |  |
| Journal |  | | *Insert the name of the journal* |  |
| Volume |  | | *Insert volume number* |  |
| Issue |  | | *Insert issue number, if available* |  |
| Page(s) |  | | *Insert page numbers, if available* |  |
| DOI |  | | https://doi.org/10.1111/jgs.14025 |  |
| Review type |  | | *Describe the type of review (e.g. scoping review, narrative review, systematic review)* |  |
| Country |  | | *List the country or countries of the review authors* |  |
| Primary contact information review |  | | *Insert email address of corresponding review author* |  |
| Review question/aim |  | | *Insert review question/aim* |  |
| Objective (if listed) |  | | *List objectives, if available. Insert new row if needed.* |  |
| Objective (if listed) |  | | *List objectives, if available. Insert new row if needed.* |  |
| Objective (if listed) |  | | *List objectives, if available. Insert new row if needed.* |  |
| Objective (if listed) |  | | *List objectives, if available. Insert new row if needed.* |  |
| Brief summary of population of interest |  | | *Summarise the population of interest and any definitions of population provided. For example: Participants aged 18 years or older in receipt of a home palliative care service, their family caregivers, or both.* |  |
| Any comparison groups? |  | | *Briefly summarise any comparison groups considered in the included studies, if applicable. For example, usual care.* |  |
| Eligibility criteria | **Inclusion** | **Exclusion** | *List or summarise any inclusion and exclusion criteria stated.   Inclusion criteria example: Inclusion criteria were: 1) must assess at least one component of palliative care; 2) must report location of care as “home of patient” irrespective of if care was delivered in an individual home, hospice, or continuing care; 3) report on any outcome; and 4) report primary data. No study designs were excluded.  State if these are not clearly specified.* |  |
| Number of databases searched |  | | *Provide count.* |  |
| Names of databases searched |  | | *List the names of databases searched.* |  |
| Month and year searched from and to |  | | *Insert the dates that the reviews span (e.g. 1950 to November 2012)* |  |
| Date of updated search, if applicable |  | | *Insert date of updated search, if applicable* |  |
| Is a full search strategy provided |  | | *Summarise whether a full search strategy is available and add a screen shot of the search strategy to the 'Search Terms' tab within this document.  For example: Example of full search strategy is present Abbreviated search strategy present Only provided key concepts of search strategy (e.g. home care) No search strategy present* |  |
| Summary of search strategy |  | | *Copy from article or summarise how search strategy was developed. For example: A scoping review was completed [13]. Quality of included studies was assessed to increase the utility of this scoping review and to add the needed quality lens to the literature. In August 2016, the following electronic databases were searched from inception: PubMed, Embase, Cumulative Index to Nursing and Allied Health Literature (CINAHL), Web of Science, Cochrane Library, EconLit, PsycINFO, Centre for Reviews and Dissemination, Database of Abstracts of Reviews of Effects, and National Health Service Economic Evaluation Database. Palliative care in the home experts were contacted to identify additional papers. Search results were limited to those published after the year 2000, to ensure that included studies were representative of modern palliative home care. Only published studies were reviewed thus ethics approval was not required. Grey literature was not included. Only English search results with human subjects were included in abstract review. Preferred reporting items for systematic reviews and meta-analyses (PRISMA) guidelines were followed to ensure methodological best practices [14]. The search strategy consisted of three concepts. First, terms for palliative care such as “palliative care,” “terminally ill,” and “end of life care” were searched. Second, terms for home care such as “home care services/trends,” “health system pathway,” and “component” were searched. Third, terms for outcomes such as “health care quality, access, and evaluation,” “quality,” and “patient satisfaction” were searched. These three concepts were combined using the Boolean operator “and.”* |  |
| Details of searching any other resources |  | | *Insert details of searching for other resources e.g. hand searches or reference lists* |  |
| Summary of analysis |  | | *Copy text summarising approach to analysis* |  |
| Number of included studies |  | | *Provide count of included studies* |  |
| Types of studies included |  | | *Describe the types of studies included (e.g. RCT (n = 1)* |  |
| Countries covered by the review |  | | *Describe the countries included in the review (e.g. UK (n = 10)* |  |
| Overall findings |  | | *Copy and paste the overall findings.* |  |
| Brief summary of intervention of interest *[To start a new line, press Alt+Enter]* |  | | *Description of intervention of interest and any definitions.   Example: A team delivering home palliative care with the presence of the following four elements:  1. Primarily for patients with a severe or advanced disease (malignant or non-malignant), no longer responding to curative/maintenance treatment or symptomatic (or both), or their family caregivers, or both. 2. Aiming to support patients or family caregivers, or both, outside hospital and other institutional settings as far as possible and to enable patients to stay at home. While conducting the review we have also included interventions in which it was clear the majority of service contacts were established while the patients were at home. Services delivered in skilled nursing facilities, day care centres, residential homes or prisons were excluded. 3. Providing either specialist or intermediate palliative/hospice care, as defined in a previous systematic review 4. Providing comprehensive care and aiming at different physical and psychosocial components of palliative care* |  |
| Primary outcome of interest |  | | *Describe the primary outcome of the review (e.g. symptom burden), add new line if needed. If not specified in advance, summarise the outcomes identified from the studies they included. For example, in Hofmeister et al. (2013):  Multiple outcomes described studies in which the objective statement identified a combination of resource use, symptom burden, quality of life, satisfaction, caregiver distress, or place of death as the primary outcome. The most commonly reported outcome was descriptive in nature with the objective of the study being to describe experiences with services offered.* ***ADD NEW ROWS IF NEEDED*** |  |
| Describe primary measures considered to assess outcome, if specified |  | | *Describe the measure used to assess the primary outcome if available, add new line if needed and may need to consult primary included studies. For example, Edmonton Symptom Assessment Scale (ESAS).* |  |
| Secondary outcome of interest |  | | *Describe any secondary outcome, add new line life needed. If not specified in advance, summarise the outcomes identified from the studies they included. For example, pain or utilisation of care.* |  |
| *Describe measure used to assess secondary outcome.* |  | | *Describe the measure used to assess the secondary outcome, add new line if needed. For example, Integrated Palliative care Outcome Scale (IPOS).* |  |
| Secondary outcome of interest |  | | *Describe any secondary outcome, add new line life needed. If not specified in advance, summarise the outcomes identified from the studies they included. For example, pain or utilisation of care.* |  |
| *Describe measure used to assess secondary outcome.* |  | | *Describe the measure used to assess the secondary outcome, add new line if needed. For example, Integrated Palliative care Outcome Scale (IPOS).* |  |
| Secondary outcome of interest |  | | *Describe any secondary outcome, add new line life needed. If not specified in advance, summarise the outcomes identified from the studies they included. For example, pain or utilisation of care.* |  |
| *Describe measure used to assess secondary outcome.* |  | | *Describe the measure used to assess the secondary outcome, add new line if needed. For example, Integrated Palliative care Outcome Scale (IPOS).* |  |
| Secondary outcome of interest |  | | *Describe any secondary outcome, add new line life needed. If not specified in advance, summarise the outcomes identified from the studies they included. For example, pain or utilisation of care.* |  |
| *Describe measures used to assess secondary outcome.* |  | | *Describe the measure used to assess the secondary outcome, add new line if needed. For example, Integrated Palliative care Outcome Scale (IPOS).* |  |
| *Model(s) of care, if identified* |  | | *Describe the model(s) of care, if specified. For this review, a ‘model of care’ has been defined as the way in which health and care services are delivered and provides ‘a descriptive picture of practice’. For example, Firth and colleagues established key criteria to define and allow for comparison between models of specialist palliative care, such as setting of care (e.g. inpatient hospital, inpatient hospice and home-based) or number of disciplines delivering care.* |  |
| Component 1 |  | | *List each of the components of care identified. For example: integrated teamwork, symptom management, holistic care, skilled providers (who are caring and compassionate), timely and responsive care, and patient and family preparedness. EACH NEEDS TO BE ENTERED ON A SEPARATE LINE. Add a new row, if needed.* |  |
| Component 2 |  | | *List each of the components of care identified. For example: integrated teamwork, symptom management, holistic care, skilled providers (who are caring and compassionate), timely and responsive care, and patient and family preparedness. EACH NEEDS TO BE ENTERED ON A SEPARATE LINE. Add a new row, if needed.* |  |
| Component 3 |  | | *List each of the components of care identified. For example: integrated teamwork, symptom management, holistic care, skilled providers (who are caring and compassionate), timely and responsive care, and patient and family preparedness. EACH NEEDS TO BE ENTERED ON A SEPARATE LINE. Add a new row, if needed.* |  |
| Component 4 |  | | *List each of the components of care identified. For example: integrated teamwork, symptom management, holistic care, skilled providers (who are caring and compassionate), timely and responsive care, and patient and family preparedness. EACH NEEDS TO BE ENTERED ON A SEPARATE LINE. Add a new row, if needed.* |  |
| Strengths |  | | *State strengths* |  |
| Limitations |  | | *State limitations* |  |
| Conclusions |  | | *Copy and paste conclusions from the review.* |  |
| Implications for practice |  | | *State implications for practice as reported by authors.* |  |
| Implications for research |  | | *State implications for research as reported by authors.* |  |
| Do the review authors need to be contacted for further information? |  | | *Please put "Yes" if there is incomplete information that is required and contact authors, update accordingly.* |  |
