## Supplemental material 1: Full search strategies for original main database searches. for "Models, components, and outcomes of palliative and end-of-life care provided to adults living at home: A systematic review of reviews"

Structure: (Palliative care AND (primary care OR community/home based care OR OOH) OR (Primary care AND OOH)) AND systematic review filter(s) if required. Major concepts are highlighted in the strategies below.

Update searches were performed in September 2022, August 2023 and August 2024 by re-running each search and limiting the results to those added to the database since the date of the last search. The final update search was performed on 01 August 2024.

Forward and backwards citation searches were performed using Citation Chaser on all included reviews following each update. The final citation search was performed on 28^th^ August 2024.

Ovid MEDLINE(R) ALL <1946 to September 22, 2021>

1	exp advance care planning/	10224

2	exp attitude to death/	16526

3	exp bereavement/	14016

4	death/	18437

5	hospices/ or "Hospice and Palliative Care Nursing"/	6643

6	life support care/	7835

7	palliative care/ or Palliative Medicine/	58128

8	exp terminal care/ or respite care/	54815

9	terminally ill/	6682

10	palliat$.af.	130902

11	hospice$.af.	37411

12	(terminal care or respite care).af.	32298

13	or/1-12	226478

14	journal of palliative care.jn.	1580

15	journal of palliative medicine.jn.	5247

16	hospice journal physical psychosocial & pastoral care of the dying.jn.	348

17	supportive care in cancer.jn.	7696

18	palliative medicine.jn.	2916

19	palliative & supportive care.jn.	1513

20	journal of supportive oncology.jn.	633

21	journal of social work in end of life & palliative care.jn.	404

22	journal of pain & symptom management.jn.	6058

23	journal of pain & palliative care pharmacotherapy.jn.	1068

24	international journal of palliative nursing.jn.	2030

25	death studies.jn.	1688

26	death education.jn.	90

27	american journal of hospice care.jn.	262

28	american journal of hospice & palliative medicine.jn.	3066

29	omega journal of death & dying.jn.	1038

30	or/14-29	35637

31	13 or 30	235928

32	bereave*.mp.	10808

33	attitude to death.mp.	16609

34	end of life.af.	28026

35	Advance* Care.af.	10913

36	((advanced or terminal*) adj (ill* or disease)).ti,ab,kw.	29655

37	supportive care.ti,ab,kw.	18115

38	dying.ti,ab,kw.	36891

39	"last year of life".ti,ab,kw.	722

40	(limited life adj (expectanc* or span*)).ti,ab,kw. or life-limiting.mp.	3642

41	or/32-40	132628

42	31 or 41 [palliative care concept]	307708

43	exp Primary Health Care/	174033

44	exp General Practice/	76472

45	family practi*.af.	88206

46	general pract*.af. or (generalist* or nonspecialist* or non-specialist*).ti,ab,kw.	137182

47	((primary adj2 care) or (primary adj2 (provider* or setting* or service*))).ti,ab,kw.	151943

48	or/43-47 [primary care concept]	417576

49	exp home care services/ or home health nursing/ or home nursing/	49050

50	((home or community) adj5 (care or nursing)).mp.	138956

51	((hospice or hospital) adj2 home).mp.	5909

52	home-based.ti,ab,kw.	12076

53	exp Community Health Nursing/ or general practitioners/ or physicians, primary care/ or Primary Care Nursing/ or ((primary or practice) adj2 nurs*).ti,ab,kw. or (general practitioner* or GP).ti,ab,kw.	144587

54	Pharmacists/ or (family adj (physician* or doctor*)).ti,ab,kw. or community paramedic*.ti,ab,kw.	38679

55	*community health services/ or community pharmacy services/ or Home Health Aides/ or Community Health Workers/ or Physical Therapists/ or Occupational Therapists/	34188

56	exp Telemedicine/ or (telemedicine or telehealth).ti,ab,kw.	46044

57	(community adj2 (care or health*)).ti,ab,kw. or community.ti,kw.	187875

58	(district adj nurs*).ti,ab,kw.	1942

59	or/49-58 [home-based or community setting concept]	514250

60	exp After-Hours Care/	2026

61	Night Care/	1393

62	(after hour* or ((outside or out or after or off) adj2 (normal or working or office) adj2 (time or hour*))).ti,ab,kw.	2711

63	after office hour*.ti,ab,kw.	48

64	(out of hours or (OOH or OOHs)).ti,ab,kw.	3686

65	out of office hours.ti,ab,kw.	55

66	(off adj hour*).ti,ab,kw.	449

67	((weekend* or evening* or holiday* or night*) adj (hour* or care*)).ti,ab,kw.	1335

68	((24 hour* or 24H or around-the-clock or around the clock) adj2 care*).ti,ab,kw.	529

69	or/60-68 [OOh concept]	10480

70	review.pt.	2863516

71	(medline or medlars or embase or pubmed or cochrane).tw,sh.	268028

72	(scisearch or psychinfo or psycinfo).tw,sh.	44844

73	(psychlit or psyclit).tw,sh.	917

74	cinahl.tw,sh.	34009

75	((hand adj2 search$) or (manual$ adj2 search$)).tw,sh.	14614

76	(electronic database$ or bibliographic database$ or computeri?ed database$ or online database$).tw,sh.	45703

77	(pooling or pooled or mantel haenszel).tw,sh.	122604

78	(peto or dersimonian or der simonian or fixed effect).tw,sh.	8752

79	(retraction of publication or retracted publication).pt.	19974

80	or/71-79	416184

81	70 and 80	177157

82	meta-analysis.pt.	142222

83	meta-analysis.sh.	142222

84	(meta-analys$ or meta analys$ or metaanalys$).tw,sh.	242514

85	(systematic$ adj5 review$).tw,sh.	249920

86	(systematic$ adj5 overview$).tw,sh.	2759

87	(quantitativ$ adj5 review$).tw,sh.	9056

88	(quantitativ$ adj5 overview$).tw,sh.	355

89	(quantitativ$ adj5 synthesis$).tw,sh.	3570

90	(methodologic$ adj5 review$).tw,sh.	7250

91	(methodologic$ adj5 overview$).tw,sh.	482

92	(integrative research review$ or research integration).tw.	155

93	((qualitativ$ adj5 synthesis$) or (scoping adj review)).tw,sh.	15078

94	Systematic Review.pt.	169097

95	or/82-94	398261

96	81 or 95 [BMJ SR filter]	463828

97	42 and (48 or 59 or 69) [Pall AND primary care or community or OOH]	29815

98	48 and 69 [Primary care and OOH]	2205

99	97 or 98	31873

100	limit 99 to (systematic reviews pre 2019 or systematic reviews)	1638 [inbuilt filter]

101	96 and 99	1262

102	100 or 101	1869

Embase <1974 to 2021 September 22>

1	advance care planning/	4033

2	attitude to death/	11271

3	bereavement/	9666

4	death/	276755

5	hospice/	14338

6	exp palliative therapy/	120991

7	respite care/	1200

8	terminal care/ or hospice care/	47165

9	exp terminally ill patient/	8972

10	palliat$.af.	199702

11	hospice$.af.	57796

12	(terminal care or respite care).af.	40481

13	supportive care.ti,ab,kw.	33536

14	bereave$.mp.	14408

15	attitude to death.mp.	11426

16	end of life.af.	40653

17	((advanced or terminal* or critical*) adj (ill* or disease)).ti,ab,kw.	132660

18	Advance* Care.af.	12717

19	(limited life adj (expectanc* or span*)).ti,ab,kw. or life-limiting.mp.	5631

20	"last year of life".ti,ab,kw.	1023

21	dying.ti,ab,kw.	47393

22	or/1-21 [palliative concept]	719997

23	exp primary health care/	183894

24	general practice/	80137

25	family practi*.af.	25876

26	general pract*.af. or (generalist* or nonspecialist* or non-specialist*).ti,ab,kw.	252972

27	((primary adj2 care) or (primary adj2 (provider* or setting* or service*))).ti,ab,kw.	204288

28	or/23-27 [primary care concept]	462300

29	exp home care/	79293

30	((home or community) adj5 (care or nursing)).mp.	249715

31	((hospice or hospital) adj2 home).mp.	8843

32	home-based.ti,ab,kw.	16663

33	exp community health nursing/	23871

34	general practitioner/ or (general practitioner* or GP).ti,ab,kw.	184323

35	((primary or practice) adj2 nurs*).ti,ab,kw.	35608

36	exp pharmacist/ or paramedical personnel/	98838

37	(family adj (physician* or doctor*)).ti,ab,kw.	25741

38	community care/ or community based rehabilitation/ or community health nursing/ or community integration/ or community mental health service/ or community program/ or preventive health service/	111275

39	"pharmacy (shop)"/	6435

40	health auxiliary/	7815

41	physiotherapist/	23183

42	occupational therapist/	7340

43	community paramedic*.ti,ab,kw.	200

44	telehealth/ or telemedicine/ or telenursing/	43158

45	(telemedicine or telehealth or telenursing).ti,ab,kw.	31193

46	(community adj2 (care or health*)).ti,ab,kw. or community.ti,kw.	223520

47	(district adj nurs*).ti,ab,kw.	1886

48	or/29-47 [home-based or community setting concept]	838128

49	out-of-hours care/	526

50	night care/	189

51	(after hour* or ((outside or out or after or off) adj2 (normal or working or office) adj2 (time or hour*))).ti,ab,kw.	4075

52	after office hour*.ti,ab,kw.	79

53	(out of hours or (OOH or OOHs)).ti,ab,kw.	5348

54	out of office hours.ti,ab,kw.	78

55	(off adj hour*).ti,ab,kw.	805

56	((weekend* or evening* or holiday* or night*) adj (hour* or care*)).ti,ab,kw.	1864

57	((24 hour* or 24H or around-the-clock or around the clock) adj2 care*).ti,ab,kw.	820

58	or/49-57 [OOH concept]	12737

59	exp review/	2829076

60	(literature adj3 review$).ti,ab.	389148

61	exp meta analysis/	226698

62	exp "systematic review"/	314005

63	or/59-62	3140183

64	(medline or medlars or embase or pubmed or cinahl or amed or psychlit or psyclit or psychinfo or psycinfo or scisearch or cochrane).ti,ab.	350843

65	RETRACTED ARTICLE/	11162

66	64 or 65	361629

67	63 and 66	278429

68	((systematic$ or scoping) adj2 (review$ or overview)).ti,ab.	299200

69	(meta?anal$ or meta anal$ or meta-anal$ or metaanal$ or metanal$).ti,ab.	276350

70	67 or 68 or 69	532221 [SR filter]

71	22 and (28 or 48 or 58) [Pall AND (primary care or community or OOH)]	54661

72	28 and 58 [primary care AND OOH]	2240

73	71 or 72	56693

74	73 and 70 [limited to SRs]	1884

75	limit 74 to conference abstract status	518

76	74 not 75	1366 [removal of conference abstracts]

The Cochrane Library

Comment:	Search for palliative care AND (community health, OOH, primary care).........and also (primary care AND OOH). No SR filter needed as just results from CDSR used

ID	Search

#1	MeSH descriptor: [Advance Care Planning] explode all trees

#2	MeSH descriptor: [Attitude to Death] explode all trees

#3	MeSH descriptor: [Bereavement] explode all trees

#4	MeSH descriptor: [Death] this term only

#5	MeSH descriptor: [Hospices] this term only

#6	MeSH descriptor: [Life Support Care] this term only

#7	MeSH descriptor: [Palliative Care] explode all trees

#8	MeSH descriptor: [Respite Care] explode all trees

#9	MeSH descriptor: [Terminal Care] explode all trees

#10	MeSH descriptor: [Terminally Ill] explode all trees

#11	(palliat* or hospice*):ti,ab,kw

#12	((terminal or supportive) next care):ti,ab,kw

#13	(respite next care):ti,ab,kw

#14	(bereave* or dying):ti,ab,kw

#15	("attitude to death"):ti,ab,kw

#16	(((advanced or terminal* or critical*) next (ill* or disease))):ti,ab,kw

#17	(advance* next care):ti,ab,kw

#18	("last year of life" or (limited next (expectanc* or span*)) or life-limiting):ti,ab,kw

#19	#1 or #2 or #3 or #4 or #5 or #6 or #7 or #8 or #9 or #10 or #11 or #12 or #13 or #14 or #15 or #16 or #17 or #18 [palliative care]

#20	MeSH descriptor: [Primary Health Care] explode all trees

#21	MeSH descriptor: [General Practice] explode all trees

#22	(family practi*):ti,ab,kw

#23	(general pract*):ti,ab,kw

#24	(generalist* or nonspecialist* or non-specialist*):ti,ab,kw

#25	(primary near/2 care):ti,ab,kw

#26	(primary near/2 (provider* or setting* or service*)):ti,ab,kw

#27	#20 or #21 or #22 or #23 or #24 or #25 or #26 [primary care]

#28	MeSH descriptor: [Home Care Services] explode all trees

#29	MeSH descriptor: [Home Health Nursing] explode all trees

#30	MeSH descriptor: [Home Nursing] explode all trees

#31	(((home or community) near/5 (care or nursing))):ti,ab,kw

#32	((hospice or hospital) near/2 home):ti,ab,kw

#33	(home-based):ti,ab,kw

#34	MeSH descriptor: [Community Health Nursing] explode all trees

#35	MeSH descriptor: [General Practitioners] explode all trees

#36	MeSH descriptor: [Physicians, Primary Care] explode all trees

#37	(((district or primary or practice) near/2 nurs*)):ti,ab,kw

#38	MeSH descriptor: [Pharmacists] explode all trees

#39	((family next (physician* or doctor*))):ti,ab,kw

#40	(general practitioner* or GP):ti,ab,kw

#41	MeSH descriptor: [Community Health Services] this term only

#42	MeSH descriptor: [Community Pharmacy Services] explode all trees

#43	MeSH descriptor: [Home Health Aides] explode all trees

#44	MeSH descriptor: [Community Health Workers] this term only

#45	MeSH descriptor: [Physical Therapists] explode all trees

#46	MeSH descriptor: [Occupational Therapists] explode all trees

#47	MeSH descriptor: [Telemedicine] explode all trees

#48	(telemedicine or telehealth or telenursing):ti,ab,kw

#49	(community near/2 (care or health*)):ti,ab,kw

#50	(community):ti

#51	(community paramedic*):ti,ab,kw

#52	{or #28-#51} [home, community care]

#53	MeSH descriptor: [After-Hours Care] explode all trees

#54	MeSH descriptor: [Night Care] explode all trees

#55	(after next hour*):ti,ab,kw

#56	(((outside or out or after or off) near/2 (normal or working or office) near/2 (time or hour*))):ti,ab,kw

#57	("after office hour*"):ti,ab,kw

#58	("(out of hours" or out-of-hours or OOH or OOHs)):ti,ab,kw

#59	((weekend* or evening* or holiday* or night*) next (hour* or care*))

#60	(((24 next hour* or 24H or around-the-clock or "around the clock") near/2 care*)):ti,ab,kw

#61	{or #53-#60} [OOH care]

#62	#19 and (#27 or #52 or #61) [Palliative care AND (primary, community, OOH]

#63	#27 and #61 [primary care AND OOH]

#64	#62 or #63

CINAHL

| **#** | **Query** | **Limiters/Expanders** | **Results** |
| --- | --- | --- | --- |
| S1 | ( (MH "Terminal Care+") or (MH "Palliative Care") or (MH "Attitude to Death") or (MH "Advance Care Planning") or (MH "Respite Care") or (MH "Hospices") or (MH "Life Support Care") ) OR TI ( bereave* or hospice* or "end of life" of "terminally ill" or palliat* ) OR AB ( bereave* or hospice* or "end of life" of "terminally ill" or palliat* ) | Search modes - Boolean/Phrase | Display |
| S2 | TI life-limiting OR AB life-limiting | Search modes - Boolean/Phrase | Display |
| S3 | TI ( ((advanced or terminal* or critical*) n1 (ill* or disease)) ) OR AB ( ((advanced or terminal* or critical*) n1 (ill* or disease)) ) | Search modes - Boolean/Phrase | Display |
| S4 | TI ( (limited life N1 (expectanc* or span*)) ) OR AB ( (limited life N1 (expectanc* or span*)) ) | Expanders - Apply equivalent subjects  Search modes - Boolean/Phrase | Display |
| S5 | S1 OR S2 OR S3 OR S4 | Expanders - Apply equivalent subjects  Search modes - Boolean/Phrase | PALLIATIVE CARE |
| S6 | (MH "Primary Health Care") | Expanders - Apply equivalent subjects  Search modes - Boolean/Phrase | Display |
| S7 | (MH "Family Practice") | Expanders - Apply equivalent subjects  Search modes - Boolean/Phrase | Display |
| S8 | TI family practi* OR AB family practi* | Expanders - Apply equivalent subjects  Search modes - Boolean/Phrase | Display |
| S9 | ( generalist* or nonspecialist* or non-specialist* ) AND ( generalist* or nonspecialist* or non-specialist* ) | Expanders - Apply equivalent subjects  Search modes - Boolean/Phrase | Display |
| S10 | TI primary N2 care OR AB primary N2 care | Expanders - Apply equivalent subjects  Search modes - Boolean/Phrase | Display |
| S11 | TI ( primary N2 (provider* or setting* or service*) ) OR AB ( primary N2 (provider* or setting* or service*) ) | Expanders - Apply equivalent subjects  Search modes - Boolean/Phrase | Display |
| S12 | S6 OR S7 OR S8 OR S9 OR S10 OR S11 | Expanders - Apply equivalent subjects  Search modes - Boolean/Phrase | PRIMARY CARE |
| S13 | (MH "Home Health Care+") OR (MH "Home Nursing") | Expanders - Apply equivalent subjects  Search modes - Boolean/Phrase | Display |
| S14 | (MH "Home Health Aides") | Expanders - Apply equivalent subjects  Search modes - Boolean/Phrase | Display |
| S15 | TI ( ((home or community) N5 (care or nursing)) ) OR AB ( ((home or community) N5 (care or nursing)) ) | Expanders - Apply equivalent subjects  Search modes - Boolean/Phrase | Display |
| S16 | TI ( ((hospice or hospital) N2 home) ) OR AB ( ((hospice or hospital) N2 home) ) | Expanders - Apply equivalent subjects  Search modes - Boolean/Phrase | Display |
| S17 | TI home-based OR AB home-based | Expanders - Apply equivalent subjects  Search modes - Boolean/Phrase | Display |
| S18 | (MH "Community Health Nursing") | Expanders - Apply equivalent subjects  Search modes - Boolean/Phrase | Display |
| S19 | (MH "Physicians, Family") | Expanders - Apply equivalent subjects  Search modes - Boolean/Phrase | Display |
| S20 | (MH "Primary Nursing") | Expanders - Apply equivalent subjects  Search modes - Boolean/Phrase | Display |
| S21 | TI ( ((district or primary or practice) N2 nurs*) ) OR AB ( ((district or primary or practice) N2 nurs*) ) | Expanders - Apply equivalent subjects  Search modes - Boolean/Phrase | Display |
| S22 | (MH "Pharmacists") | Expanders - Apply equivalent subjects  Search modes - Boolean/Phrase | Display |
| S23 | TI ( (family n1 (physician* or doctor*)) ) OR AB ( (family n1 (physician* or doctor*)) ) | Expanders - Apply equivalent subjects  Search modes - Boolean/Phrase | Display |
| S24 | (MH "Community Health Services") | Expanders - Apply equivalent subjects  Search modes - Boolean/Phrase | Display |
| S25 | (MH "Home Health Aides") OR (MH "Community Medicine") | Expanders - Apply equivalent subjects  Search modes - Boolean/Phrase | Display |
| S26 | (MH "Physical Therapists") | Expanders - Apply equivalent subjects  Search modes - Boolean/Phrase | Display |
| S27 | (MH "Occupational Therapists") | Search modes - Boolean/Phrase | Display |
| S28 | TI community paramedic* OR AB community paramedic* | Search modes - Boolean/Phrase | Display |
| S29 | (MH "Telehealth") OR (MH "Telemedicine+") | Search modes - Boolean/Phrase | Display |
| S30 | TI ( telemedicine or telehealth or telecare ) OR AB ( telemedicine or telehealth or telecare ) | Search modes - Boolean/Phrase | Display |
| S31 | TI community OR AB ( community N2 (care or health*) ) OR TI ( community N2 (care or health*) ) | Search modes - Boolean/Phrase | Display |
| S32 | TI district N1 nurs* OR AB district N1 nurs* | Search modes - Boolean/Phrase | Display |
| S33 | S13 OR S14 OR S15 OR S16 OR S17 OR S18 OR S19 OR S20 OR S21 OR S22 OR S23 OR S24 OR S25 OR S26 OR S27 OR S28 OR S29 OR S30 OR S31 OR S32 | Search modes - Boolean/Phrase | HOME/COMMUNITY CARE |
| S34 | TI out of hours | Search modes - Boolean/Phrase | Display |
| S35 | (MH "Night Care") | Search modes - Boolean/Phrase | Display |
| S36 | TI ( ((outside or out or after or off) N2 (normal or working or office) N2 (time or hour*))) ) OR AB ( ((outside or out or after or off) N2 (normal or working or office) N2 (time or hour*))) ) | Search modes - Boolean/Phrase | Display |
| S37 | TI "after hour*" OR AB "after hour*" | Search modes - Boolean/Phrase | Display |
| S38 | TI out of hour* OR AB out of hour* | Search modes - Boolean/Phrase | Display |
| S39 | TI "off hour*" OR AB "off hour*" OR TI ( ((weekend* or evening* or holiday* or night*) N1 (hour* or care*)) ) OR AB ( ((weekend* or evening* or holiday* or night*) N1 (hour* or care*)) ) | Search modes - Boolean/Phrase | Display |
| S40 | TI ( ((24 hour* or 24H or around-the-clock or around the clock) N2 care*) ) OR AB ( ((24 hour* or 24H or around-the-clock or around the clock) N2 care*) ) | Search modes - Boolean/Phrase | Display |
| S41 | S34 OR S35 OR S36 OR S37 OR S38 OR S39 OR S40 | Expanders - Apply equivalent subjects  Search modes - Boolean/Phrase | OUT OF HOURS |
| S42 | S12 or S33 OR S41 | Expanders - Apply equivalent subjects  Search modes - Boolean/Phrase | Primary, home, community or OOH |
| S43 | S5 AND S42 | Expanders - Apply equivalent subjects  Search modes - Boolean/Phrase | Palliative AND (primary, home, community or OOH) |
| S44 | S12 AND S41 | Expanders - Apply equivalent subjects  Search modes - Boolean/Phrase | Primary care and OOH |
| S45 | S43 OR S44 | Expanders - Apply equivalent subjects  Search modes - Boolean/Phrase | Palliative AND (primary, home, community or OOH)  OR  Primary care and OOH |
| S46 | (TI (systematic* n3 review*)) or (AB (systematic* n3 review*)) or (TI (systematic* n3 bibliographic*)) or (AB (systematic* n3 bibliographic*)) or (TI (scoping n3 review)) or (AB (scoping n3 review)) or (TI (systematic* n3 literature)) or (AB (systematic* n3 literature)) or (TI (comprehensive* n3 literature)) or (AB (comprehensive* n3 literature)) or (TI (comprehensive* n3 bibliographic*)) or (AB (comprehensive* n3 bibliographic*)) or (TI (integrative n3 review)) or (AB (integrative n3 review)) or (JN “Cochrane Database of Systematic Reviews”) or (TI (information n2 synthesis)) or (TI (data n2 synthesis)) or (AB (information n2 synthesis)) or (AB (data n2 synthesis)) or (TI (data n2 extract*)) or (AB (data n2 extract*)) or (TI (medline or pubmed or psyclit or cinahl or (psycinfo not “psycinfo database”) or “web of science” or scopus or embase)) or (AB (medline or pubmed or psyclit or cinahl or (psycinfo not “psycinfo database”) or “web of science” or scopus or embase)) or (MH “Systematic Review”) or (MH “Meta Analysis”) or (TI (meta-analy* or metaanaly*)) or (AB (meta-analy* or metaanaly*)) | Search modes - Boolean/Phrase | SR filter |
| S47 | S45 AND S46 | Expanders - Apply equivalent subjects  Search modes - Boolean/Phrase | Display |

Epistemonikos

(title:(palliat* OR hospice* OR "end of life" or “last year of life”) OR abstract:(palliat* OR hospice* OR "end of life" or “last year of life”)) AND (title:(community OR "primary care" OR "general practic*" OR "family practice" OR "district nurs*" OR home-based OR "at home" or “in the home” or “in-home” or “in home”OR "out of hours" OR OOH OR OOHs OR "after hours") OR abstract:(community OR "primary care" OR "general practic*" OR "family practice" OR "district nurs*" OR home-based OR "at home" OR "out of hours" OR OOH OR OOHs OR "after hours"))
