## Supplemental material 2: Characteristics of included reviews. for "Models, components, and outcomes of palliative and end-of-life care provided to adults living at home: A systematic review of reviews"

| **Author & country** | **Type of review** | **Time period of the search** | **Type and number of studies included** | **Population (number)** | **Interventions** | **Primary outcomes** | **Quality appraisal in study (Yes/ No & tool)** | **Study quality (AMSTAR= grading or SANRA= score)** |
| --- | --- | --- | --- | --- | --- | --- | --- | --- |
| Ahn et al., 2020 ^40^ | Systematic review | 2007-2018 | n= 11 | n= 1361 | Caregiver support intervention | Psychological distress, caregiving burden, quality of life, self-efficacy, and competence for caregiving. | Yes | AMSTAR - 2 rating: Critically low |
|  |  |  | RCTs (n=9), Quasi-experimental study (n=2) |  |  |  | Cochrane Risk of Bias (RoB) tool for RCTs and ROBINS-I |  |
| United States of America |  |  |  | Caregivers of patients with advanced cancer (Stage III or IV) |  |  |  |  |
| Aoun et al., 2014^85^ | Narrative review | 2002-2013 | n= 37 | n= not provided | Care aid support & Personal alarm use | Place of care & death & physical and psychological wellbeing. | No | SANRA Score= 11 |
| Australia |  |  | Retrospective (n= 21), NRCT (n= 1), Longitudinal (n=2), Qualitative (n= 4), Prospective (n= 8), cross-sectional (n= 1) | Terminally ill adults who live alone at home, adult caregivers of people who died 9 months earlier, healthcare professionals caring for these patients |  |  |  |  |
| Bainbridge et al., 2016 ^11^ | Review of systematic reviews | 1950-2014 | n= 30 | n= not provided | Multidisciplinary Specialist palliative care in-home services; Multidisciplinary Palliative care hospital outreach teams | Quality of life (QoL), care satisfaction, individual performance status, pain management, supporting home death, non-pain symptom management. | No | AMSTAR - 2 rating: Critically low |
| Canada |  |  | Systematic reviews (n=17), narrative review (n=2), RCT (n= 13), Quasi- experimental study (n=4), Cohort (n= 15), Pre/post (n=8) | Home treatment programs for people with an advanced or palliative illness. |  |  |  |  |
| Basile et al., 2023 ^41^    Italy | Scoping review | NA | n= 14  Quantitative (n= 8), Qualitative (n= 2), Mixed methods (n=4) | n= not provided | Not specified by paper, but under model of telehealth | Process of care, assessment of patient needs, goal setting, care plans, outcome monitoring, intervention reporting frequency, communication effectiveness, and patient perspectives on positive or negative aspects of technology. | No | Adhered to the Preferred Reporting Items for Systematic Reviews and Meta-analyses Extension for Scoping Reviews (PRISMA-ScR) checklist |
|  |  |  |  | Adults aged over 65 in receipt of palliative and/or end-of-life care at home and using technology. |  |  |  |  |
| Bayly et al., 2021 ^42^ | Tertiary systematic review | 2000-2019 | n= 78 | n= 17,739 | Geriatric Integrated Care AND Palliative Integrated Care | QoL, Utilisation of acute services or community services, Costs of health services utilisation, | Yes | AMSTAR - 2 rating: Low |
| United Kingdom |  |  | Review-level literature that can include trials that are randomised (cluster, parallel, single stage or cross-over design), nonrandomised trials, controlled before after studies, interrupted time series studies and repeated measures studies. | Older people aged 60 years and over with advanced progressive health conditions. |  |  | A Measurement Tool to Assess systematic Reviews (AMSTAR) |  |
| Brereton et al., 2017 ^10^ | Review of systematic & narrative reviews | 2000-2014 | n= 13, systematic & narrative reviews | n= not provided | 1. Home-based palliative care (i.e. models of palliative care delivered within the patient or their carer’s own home); 2. Models delivered across multiple settings. 3. Palliative care approach (including outpatient palliative care. 4. Palliative care teams. | Patient outcomes and family or caregiver outcomes. Patient and caregiver outcomes relate to physical (e.g. physical symptoms), psychological (e.g. coping) and social issues (e.g. place of care) | Yes | AMSTAR - 2 rating: Critically low |
|  |  |  |  | Reviews considering adults (defined as people aged 18 and over) with life-limiting illnesses as defined by the study authors. |  |  | A Measurement Tool to Assess systematic Reviews (AMSTAR) |  |
| United Kingdom |  |  |  |  |  |  |  |  |
| Carmont et al., 2018 ^86^ | Systematic review | Not stated | n= 17, | n= not provided | Integrated primary and secondary care delivering care at home. | Performance status, pain intensity, Hospitalisation, QoL, symptoms, time to death or readmission, all cause hospital bed days, ED visits, ED Admissions that then led to later discharge, Recommendations followed in care plan, Perceived usefulness of case, Opinions of community healthcare providers regarding palliative care service, Qualitative analysis of GP lead case conference. | No | AMSTAR - 2 rating: Low |
| Australia |  |  | RCT (n=3), Cohort (n=1), Surveys (n=4), Qualitative (n=7), Narrative (n=2) | Adults receiving care from their GP, specialist hospital services or an integrated model of care. |  |  |  |  |
| Chen et al., 2022 ^43^ | Systematic review | Up to 2020 | n= 5 | n= 3936 | Home health care in comparison to alternative palliative care | Advance care planning, place of death, Healthcare utilisation. | Yes | AMSTAR - 2 rating: Critically low |
| United Kingdom, Taiwan, The Netherlands, Belgium |  |  | Prospective cohort study (n=1), retrospective cohort study (n=2), Case control studies (n=2) | Adults receiving home health care |  |  | The Critical Appraisal Skills Programme (CASP) toolkit |  |
| Chen et al., 2023 ^44^ | Systematic review | Up to 2021 | n= 30 | n= not provided | Telehealth palliative care interventions | Quality of life, burden, survival, mood, advance care planning (ACP), utilisation, satisfaction | Yes | AMSTAR - 2 rating: Moderate |
| China |  |  | RCT (n=30) | Adult patients (aged 18 and older) with life-limiting illness (defined by classifications of disease severity) |  |  | Cochrane Collaboration’s tool. |  |
| Critchley et al., 1999 ^97^ | Systematic review | From 1966 to 1997 | n= 39 | n= not provided | Hospice care model, home care model, home based palliative care nursing service model, hospice at home model. | Health service outcomes, patient outcomes and family or caregiver outcomes. Patient and caregiver outcomes relate to physical (e.g. physical symptoms), psychological (e.g. coping) and social issues (e.g. place of care). | Yes | AMSTAR - 2 rating: Critically low |
|  |  |  | Eleven studies were RCTs; 30 non-randomized comparative studies. | Adults receiving palliative care |  |  | Jadad Scale |  |
| Canada |  |  |  |  |  |  |  |  |
| Davies and Higginson., 2005 ^98^ | Systematic review | Up to 2003 | n= 15 | Adults with cancer attending day care facilities | Specialist palliative day care services | Symptom control, quality of life, social and psychological support, and patient and relative satisfaction with care | Yes | AMSTAR - 2 rating: Critically low |
|  |  |  | 15 papers report on data from 12 observational studies. Of which, 11 studies were UK-based and one was from the United States of America. |  |  |  | Author-developed scale |  |
| United Kingdom |  |  |  |  |  |  |  |  |
| Davis et al., 2015 ^87^ | Systematic review | Not provided | n= 20 | n= not provided | Earlier palliative care integration at home and as an outpatient. | Symptoms, QoL, hospitalisations, cost, caregiver quality of life, caregiver burden | No | AMSTAR - 2 rating: Critically low |
| United States of America |  |  | Fifteen RCTs of outpatient palliative care and 13 RCTs of palliative home care, 7 SRs | Patients with a serious illness as an outpatient and at home. |  |  |  |  |
| de Nooijer et al., 2020 ^45^ | Systematic review | Up to 2019 | n= 10 | n= not provided | Specialist palliative care | Symptoms, quality of life, satisfaction with care, hospitalisation, place of death | Yes | AMSTAR - 2 rating: Low |
| Belgium |  |  | Qualitative design (n=3), Quantitative design (n=3), Mixed Methods (n=3) Narrative review (n=1) | Studies concerning older people in the primary/ community care setting using palliative care services |  |  | Qualitative and quantitative assessment scales |  |
| DeGroot et al., 2020 ^46^ | Integrative review | Up to 2019 | n= 19 | n= not provided | A team delivering palliative care to patients with advanced heart failure or supporting their caregivers | Symptom, function status, QoL, symptom burden, advanced care planning, hospitalisation | Yes | AMSTAR - 2 rating: Critically low |
| United States of America |  |  | Quantitative (n=6); RCT (n=6); Qualitative (n=6); mixed-methods (n= 1) | Patients with palliative care receiving out-patient, home based or community based palliative care, their professionals or caregivers |  |  | Johns Hopkins Nursing Evidence-Based Practice Appraisal tools |  |
| Disalvo et al., 2021 ^47^ | Systematic review of reviews 'meta-review' | 1997- 2019 | n= 16 | n= 31718 (incl. patients/ caregivers/ HCPs) | Digital health technologies | Symptom management/reduction, Patient anxiety/depression/ psychological wellbeing, Pain assessment/management, Caregiver quality of life -Caregiver mood/anxiety - Caregiver perception of pain management, Physical function - Caregiver burden, Family functioning, Hospital/ED admissions and/or readmissions, | Yes | AMSTAR - 2 rating: Moderate |
|  |  |  | Integrative review = 2, systematic scoping review = 2, systematic review = 9, rapid review of systematic reviews = 1, systematic mixed-methods review = 1, systematic review of systematic reviews = 1 | People with palliative care needs and/or their carers at home (All ages/no restrictions on age) |  |  | Risk of Bias in Systematic Reviews (ROBIS) phase 2 domains and phase 3 |  |
| Australia |  |  |  |  |  | Patient and/or carer perceptions/user satisfaction/ satisfaction with care, experience measures, Cost/cost-effectiveness, Efficacy/effectiveness, Patient acceptance/compliance of intervention |  |  |
| Ebneter et al., 2022 ^48^ | Scoping review | 2010 onward | n= 27 | n= not provided | Telemedicine intervention | Feasibility, acceptability and needs of patient | Yes | Adhered to the Preferred Reporting Items for Systematic Reviews and Meta-analyses Extension for Scoping Reviews (PRISMA-ScR) checklist |
| Switzerland |  |  | Systematic review n = 3 (completed), n= 3 (ongoing); RCT n=1; Cohort study n=2; Cross-sectional survey n=2; Non-randomised trial n=1; Single-arm intervention study n=9; Qualitative study n= 4; Review (other) n =5; other n=2; pilot study n =1 (ongoing) | Adult in/outpatient palliative care patient/caregivers |  |  | Physician Data Query (PDQ®) level of evidence developed by the National Cancer Institute specifically for studies in the field of supportive and palliative care |  |
| Fasolino et al., 2023 ^49^ | Rapid review | January 2020 and January 2023 | n=22 | n = not provided | Telehealth for palliative services and hospice care in rural communities | Assessed outcomes at 3 levels (person- and caregiver-centred, provided-centred, and organizational-centred) | No | AMSTAR - 2 rating: Critically low |
|  |  |  | RCT 3, Literature Synthesis 1, Protocol 1, Rapid Review 1, Mixed-methods, 1, Protocol development/community engagement 1, Quantitative 1, Retrospective review 1, Systematic review 1, Efficacy 1, Longitudinal 1, Qualitative 1, Best Practice Compilation 1 | Participants in the rural US in rural communities who in receipt of telemedicine for palliative care and hospice services. |  |  |  |  |
| United States of America |  |  |  |  |  |  |  |  |
| Feliciano et al., 2024 ^50^ | Systematic review | 2013-2023 | n=9 | n= 27552 | Home-based Palliative Care | Quality of life (QoL), symptom control/burden, healthcare resource utilization (hospital admissions, emergency department visits (EDV), and length of hospital stay (LHS)) and place of death. | Yes | AMSTAR - 2 rating: Critically Low |
|  |  |  |  | Adults (≥18 years old) of any gender diagnosed with severe, prolonged, or progressive illnesses requiring end-of-life care |  |  | The revised Cochrane Risk of Bias (RoB) tool for RCTs and ROBINS-I assessment tool (cohort-type version) |  |
| Portugal |  |  | RCTs (n=5), Retrospective cohort study (n=3), Retrospective population-based study (n=1) |  |  |  |  |  |
| Finlay et al., 2002 ^99^ | Systematic review | Not provided | Not reported | n= 43 | Palliative Care Team | Other symptoms, Quality of life Satisfaction, Referral to other services Therapeutic, interventions, Carer: satisfaction Burden/morbidity, Carer Satisfaction, death rates, Health service use and costs | No | AMSTAR - 2 rating: Critically low |
| United Kingdom |  |  |  | Palliative patients |  |  |  |  |
| Finucane et al., 2021 ^51^ | Systematic review of published systematic reviews (i.e. a meta-review) | 2006 - 2018 | n=21 | n = not provided | Digital health interventions (DHI) | No restrictions were placed on outcomes as potentially influenced by palliative care DHIs. | Yes | AMSTAR - 2 rating: Moderate |
|  |  |  | Case study (8) Cost analysis (3) Cross-sectional (4) Descriptive (16) Expert opinion (1) Feasibility study (10) Interrupted time series (2) Mixed methods (41) Non-experimental (1) Non-randomised controlled trial (1) Pre- and Post- Interventions (26) Prospective cohort (5) Protocols (6) Qualitative (52) Quantitative (9) Quasi-experimental (5) Questionnaire/online survey (19) Randomised inferiority study (1) Rapid evaluation (1) RCTs (25) Realist evaluation (1) Record review (2) Retrospective cohort (8) Reviews (4) Secondary analysis (4) Service evaluation (5) System report (1) | Terminally ill patients and their families. |  |  | AMSTAR - 2 and Physician Data Query (PDQ) level of evidence |  |
| United Kingdom |  |  |  |  |  |  |  |  |
| Firth et al., 2023 ^24^ | Mixed method systematic narrative review | 1st January 1990 to 1st August 2022 | n= 64 | n= not provided | Not specified - 24-hour access to specialist palliative care; non-specialist palliative care. | Preferred place of death, physical and mental functioning, Emergency department use and economic evaluation | Yes | SANRA Score= 12 |
| United Kingdom, Taiwan |  |  | Quantitative studies (n=38) (6 RCT’s, 5 controlled cohort, 24 observational and 3 pilot studies), Qualitative studies (n=14), Mixed methods (n=4), and Service development papers (n=8) | Adults (over 18 years) with advanced illness from malignant or non-malignant disease in the last year of life or their family caregivers receiving out-of-hours palliative care intervention or service. |  |  | **For qualitative/ quantitative or both**: Quality Assessment Criteria for Evaluating Primary Research Papers (QualSyst) **For quality improvement studies:** Quality Improvement Minimum Quality Criteria Set (QI-MQCS) |  |
| Fulton et al., 2019 ^52^ | Systematic review and meta-analysis | Inception to November 2016 | n= 10 | n= 2385 | Integrated palliative outpatient care | QoL, survival, healthcare utilisation, symptom burden, psychological burden, end of life care outcomes, caregiver outcomes | Yes | AMSTAR - 2 rating: Low |
| United States of America |  |  | 8 RCTs and 2 cluster RCTs | Advanced cancer patients using integrated palliative and oncology service and their carers |  |  | Cochrane risk of bias tool for randomized controlled trials (RCTs) and the revised Newcastle—Ottawa Scale for cohort studies. Individual studies were assigned a summary risk of bias score (low, moderate, or high) |  |
| Gomes et al., 2013 ^13^ | Systematic review | Inception to November 2012 | n= 23 | n= 37561 | Home based palliative care | Symptoms, quality of life, advanced care planning, death at home, satisfaction, hospital cost | Yes | AMSTAR - 2 rating: High |
|  |  |  |  |  |  |  | Cochrane Effective Practice and Organisation of Care (EPOC) Review Group for RCTs/CCTs, CBAs and ITSs checklist |  |
| United Kingdom |  |  | 16 RCTs, 4 Controlled Clinical Trials (CCTs), 2 Controlled Before-After Studies (CBAs), 1 Interrupted Time Series (ITS) with nested CBA. | Patients receiving palliative care at home |  |  |  |  |
| Gonzalez-Jaramillo et al., 2021 ^53^ | Systematic review | 2013 to 11 February 2019 | n= 19 | n= 92000 | Home based palliative care | Hospital use, healthcare cost, place of death | Yes | AMSTAR - 2 rating: Low |
| United Kingdom |  |  | 12 retrospective cohort studies, 5 quasi-experimental studies, 2 RCTs | Adults aged greater than 18, at the end of life, with a severe illness or a disease end-stage. |  |  | Joanna Briggs Institute Critical Appraisal Tools Checklists for use in systematic reviews. |  |
| Goodrich et al., 2024 ^54^ | Additional analysis of systematic review evidence | 1st January, 1990 to 1st August 2022 | n=31 | n = not provided | Out-of-hours services | Service utilisation of the community-based palliative care services out-of-hours. This includes: Time of contact, Who contacted, Type of contact, and background of staff delivering the out-of-hours care. | Yes | SANRA Score= 12 |
|  |  |  | Retrospective review (n=12), Prospective non-randomised study (n=2), Quasi experimental (n=1), Practice development report (n= 2), RCT (n=1), Qualitative study (n=2), Service evaluation (n=11) | Palliative care patients and their families living in the community setting for example, their own homes |  |  | **For qualitative/ quantitative or both**: Quality Assessment Criteria for Evaluating Primary Research Papers (QualSyst) **For quality improvement studies:** Quality Improvement Minimum Quality Criteria Set (QI-MQCS) |  |
| United Kingdom |  |  |  |  |  |  |  |  |
| Gordon et al., 2022 ^55^ | Rapid review | 2004-2019 | n= 18 | Patients n= 3,213 Healthcare providers n= 250 | Telehealth interventions | Symptom, satisfaction, quality of life | Yes | AMSTAR - 2 rating: Critically low |
| United States of America |  |  | RCT = 1, Explorative = 1, Mixed methods = 7, Descriptive = 6, retrospective chart study = 1, clinical trial = 2 |  |  |  | Clinical Practice Guidelines for Quality Palliative Care developed by the National Coalition for Hospice and Palliative Care. |  |
| Hancock et al., 2019 ^56^ | Systematic review | After 2010 | n= 30 | n= not provided | Telehealth initiative in the delivery of palliative care in the UK. | Patient and caregiver satisfaction, number of acute hospital admission, length admission. | Yes | AMSTAR - 2 rating: Critically low |
| United Kingdom |  |  | Qualitative (7), service evaluations (4), RCT (3), protocols (3), intervention without any identifiable study design (3), randomised crossover trial (1), mixed methods (2), realist evaluation (1), prospective interventional (1), prospective longitudinal cohort (1), prospective observational (2) and retrospective observational (2) | Palliative care patients receiving telehealth interventions. |  |  | Criteria adapted from Wallace et al.’s 2004 paper on meeting the challenge of developing systematic |  |
| Hayes Bauer et al., 2024 ^57^ | Systematic integrative review | January 2011 - February 2023 | n=44 | n = not provided | Telepalliative care | Patient and family perspective and effectiveness of the Telepalliative care model. | Yes | AMSTAR - 2 rating: Critically Low |
|  |  |  | Qualitative (n=19), Quantitative (n=5), and Mixed methods (n=20) | Adult patients 18+ years and their family, engaged in Telepalliative care including generalist and specialist palliative care |  |  | Mixed Methods Approval Tool (MMAT) version 2018 |  |
| Denmark, Australia, Germany |  |  |  |  |  |  |  |  |
| Head et al., 2017 ^88^ | Systematic review | 2006-2016 | n= 11 | n= not provided | An intervention delivered to patients with end of life or palliative care, using telemedicine or telehealth intervention | Symptoms, quality of life, psychological distress, hospitalisation and cost, patient satisfaction | Yes | AMSTAR - 2 rating: Critically low |
| United States of America |  |  | Case report (n=4); non-randomized interventional n=1, randomized non-inferiority n =1, RCT n=1; mixed methods n=2; Survey n=2 | Patients receiving palliative or end -of-life care for a serious condition (i.e. advanced disease, end-stage disease) |  |  | Using the Cochrane Collaboration’s tool for assessing risk of bias |  |
| Hofmeister et al., 2018 ^89^ | Scoping review | After 2000 | n= 53 | n= not provided | Home based palliative care | Resource use, symptom burden, quality of life, satisfaction, caregiver distress, or place of death as the primary outcome Resource use, symptom burden, quality of life, satisfaction, caregiver distress, place of death, cost analysis, or described experiences. | Yes | AMSTAR - 2 rating: Low   Study was conducted using the PRISMA 2009 statement |
| Canada |  |  | Case-control = 1, cross-sectional = 4, cost analysis = 7, qualitative = 10, RCT = 7, cohort = 14, quasi experimental = 4, pre-post design = 6 | Terminally ill patients at the end of life being care for in the home. |  |  | Critical Appraisal Skills Programme Qualitative Checklist |  |
| Hughes et al., 2023 ^58^ | Systematic review | Inception to August 2021 | n=57 | n=48 (Qualitative study only) | Community-based palliative care | Effectiveness of the interventions using the following outcomes from the included studies: Advance care planning, Costs, Death location, Hospice utilisation, Hospital utilisation, Knowledge, Quality of life, Symptoms | Yes | AMSTAR - 2 rating: Critically Low |
|  |  |  | Quantitative (n=25), Qualitative (n=21), and Mixed method (n=11) research articles | Key and vulnerable population such as Rural, Lower-income, Marginalised racial or ethnic groups |  |  | Mixed Methods Approval Tool (MMAT) version 2018 |  |
| United States of America |  |  |  |  |  |  |  |  |
| Janke et al., 2024 ^59^ | Systematic review | inception to 26th January 2023. | n=7 | n= 805 | Palliative care for non-cancer patient groups | Health care costs such as hospital costs, costs borne by patients, or costs borne by patients’ family members | Yes | AMSTAR - 2 rating: Critically Low |
|  |  |  | RCTs | Adult patients (≥ 18 years) with non-cancer life limiting illnesses from the following non-cancer disease groups used in the Global Atlas of Palliative Care: lung diseases, heart diseases, cerebrovascular diseases, central nervous system diseases, diseases of liver, renal failure, HIV and dementia |  |  | Drummond's checklist |  |
| United Kingdom, India |  |  |  |  |  |  |  |  |
| Johansson et al., 2024 ^60^ | A rapid systematic review | Inception to February 9, 2023 | n=21 | n=22517 (Participants/data source) | Out-of-hours palliative care telephone advice lines | Patient/carer outcomes (health status, wellbeing and coping), and health care system outcomes (impact on the use of care services).  Cost-effectiveness was differentiated as a separate outcome while patient satisfaction and experience were incorporated as facets of patients/carer outcome. | Yes | AMSTAR - 2 rating: Low |
|  |  |  | Quantitative (n=8), Mixed methods (n=5), Qualitative (n=1), Others (n=7) | Adults (i.e. aged ≥18 years) living at home with palliative care needs and family/unpaid carers |  |  | Mixed Methods Approval Tool (MMAT) version 2018 |  |
| United Kingdom |  |  |  |  |  |  |  |  |
| Johnson et al., 2024 ^61^ | Systematic review with meta-analysis and meta-regression | January 1, 2000 to December 28, 2023 | n = 39 | n=6089 | Specialist palliative care (SPC) | Quality of life, and Emotional wellbeing | Yes | AMSTAR - 2 rating: High |
|  |  |  | RCTS (n=39) | Adults (18+) with advanced illness with palliative care needs. |  |  | Cochrane Risk of Bias 2 tool |  |
| United Kingdom, Switzerland |  |  |  |  |  |  |  |  |
| Johnston et al., 2020 ^16^ | Systematic review | 2000 - 2019 | n= 0 | n= 0 | Home based palliative care | Quality of life, survival, cost | Yes | AMSTAR - 2 rating: Low |
| Ireland |  |  | The review found no studies evaluating the effectiveness or cost-effectiveness of out-of-hours palliative care. | Adults (>18 years old) in the last year of life/ with a terminal illness /had other serious medical needs, or are carer for someone with these needs |  |  | Critical Appraisal Skills Programme (CASP) |  |
| Kidd et al., 2010 ^95^ | Narrative review | 1999-2009 | n= 21 | n= not provided | Adults or children with palliative care needs, their relatives and carers, or health professionals using telehealth, telemedicine or Information technology within the UK | Three main outcomes of interest were reported  1) Who is using telehealth 2) what is telehealth being used for 3) is telehealth use increasing | No | SANRA Score= 9 |
| United Kingdom |  |  | descriptive (n=10); pilot study audit (n=1); pilot feasibility study - exploratory design (n=2); web-based resource (n=1); mixed methods (n=3); prospective cohort (n=1); (qualitative n=1); case study (n=1); | Adults or children with palliative care needs, their relatives and carers, or health professionals using telehealth |  |  |  |  |
| Kirtania et al., 2023 ^62^ | Rapid review | Inception to June 2022 | n=7 | n = not provided | Home-based Palliative Care | For the Indian context: a) most effective and essential elements necessary for a home-based palliative care; b) challenges and experiences faced by patients/caregivers and healthcare providers; c) important and evident strategies for implementing a home-based palliative care intervention | No | AMSTAR - 2 rating: Critically Low |
|  |  |  | Mixed method systematic Review (n=1), Qualitative (n=3), Descriptive (n=1), Integrative review (n=1), Retrospective (n=1), | Patient’s and caregiver’s receiving palliative care at home |  |  |  |  |
| India |  |  |  |  |  |  |  |  |
| Layne et al., 2024 ^63^ | Integrative review | Inception to May 2023 |  | n=9237 | Care coordination | Service delivery, leadership and governance, workforce, financing, technologies and medical products, information and research reviewed at the micro, meso, and macro levels | Yes | AMSTAR - 2 rating: Critically Low |
|  |  |  | RCT (n=4), Qualitative (n=2), Mixed methods (n=3), Cohort (n=2), Case report (n=1), Program descriptions (n=3), and Study protocols (n=2) | Community-dwelling persons living with Alzheimer’s Disease and Related Dementias (ADRDs) and their caregivers. Informal caregivers were defined as spouses, parents, relatives, or friends. |  |  | Mixed Methods Appraisal Tool (MMAT) Version 2018 |  |
| United States of America |  |  |  |  |  |  |  |  |
| Luckett et al., 2013 ^15^ | Systematic review and meta-analysis | From 2011 | n= 9 | n= not provided | Specialist palliative care provided at home | Symptoms, QoL, place of death, cost | Yes | AMSTAR - 2 rating: Critically low |
| Australia |  |  | Non-randomised study (n=2); Retrospective record review n=5, RCT n=2 | People with life limiting illnesses being nursed exclusively in the home environment. Chronic illness was not included in the definition of life limiting illnesses |  |  | Study quality was independently rated using Cochrane grades |  |
| Lupati et al., 2023 ^64^ | Systematic review | January 2012 - September 2019. | n=30 |  | Community based specialist palliative care interventions | 1. Patient-related outcomes, for example, quality-of-life measures, symptom intensity, and survival. 2. Caregiver-related outcomes, for example, caregivers’ burden score. 3. Equity measures—outcomes specific to disadvantaged or Indigenous patient groups, for example, cultural appropriateness of services, uptake of services by disadvantaged or Indigenous groups. 4. Integration of specialist with non-specialist palliative care services, for example, uptake of primary care services in delivering aspects of palliative care such as end-of-life care.  5. Utilization of hospital services, for example, hospitalization, visits to emergency department (ED). | Yes | AMSTAR - 2 rating: Low |
|  |  |  | 12 observational studies, 5 RCTs, 5 qualitative studies, 8 systematic reviews | Patients, carers receiving care, or non-specialist palliative care providers (e.g., primary care) accessing support services from community-based specialist palliative care providers. Studies of Indigenous populations were considered part of the main analysis and synthesis |  |  | AMSTAR checklist, Cochrane tool for assessing risk of bias (RoB 2), ROBINS-I and CASP checklist |  |
| New Zealand |  |  |  |  |  |  |  |  |
| Luta et al., 2021 ^65^ | Narrative review of reviews | January 2000 to September 24th 2019 | n= 43 | n= not provided | Any interventions in palliative care for palliative care groups. | Economic outcomes including cost | Yes | SANRA Score= 12 |
| Switzerland and United Kingdom |  |  | Systematic reviews | Reviews considering terminally ill adults (18 years old and over) and considering patients with varying illnesses, in receipt of palliative interventions |  |  | Using the AMSTAR - 2 tool |  |
| Marshall et al., 2023 ^66^ | Systematic review | January 1, 1990 to May 2022 | n=14 | n=1831 | All novel model of care interventions | Outcomes reported using empirical data. These include: rate of death at patient preferred place, improved care, rate of hospital use, quality of life and mood, increased Kaplan-Meier 1-year survival rates, reduced hospital usage/admissions/length of stay, Symptom, hospital resource usage, functional decline, healthcare personnel and general practitioner visits. | Yes | AMSTAR - 2 rating: Low |
|  |  |  | Mixed method evaluation (n=1), Quasi-experimental (n=2), Retrospective cohort (n=1), Prospective longitudinal (n=2), Prospective mixed method (n=1), Retrospective Patient record audit (n=1), Cross sectional (n=1), RCT -Phase 3 (n=1), Case series with pre-post-test (n=1), RCT - Phase 2 (n=1), Comparative (n=1) Pseudorandomised control study (n=1) | Children and adults receiving palliative/end of life care; all health care personnel and non-health care personnel who contribute to the delivery of palliative/end of life care; whom are located in a rural setting of a high-income country |  |  | Critical Appraisal Skills Programme (CASP) checklists |  |
| Australia |  |  |  |  |  |  |  |  |
| Matthews et al., 2023 ^67^ | Scoping review | inception to June 2020 | n=23 | n=4016 | Telehealth | Patient or caregiver outcomes. These include: physical and psychological symptoms, quality of life, and acceptability or satisfaction, survival, usage patterns, feasibility, healthcare utilization and cost. | Yes | Adhered to the Preferred Reporting Items for Systematic Reviews and Meta-analyses Extension for Scoping Reviews (PRISMA-ScR) checklist |
| Canada |  |  | 7 randomized controlled trials, 5 feasibility trials, 3 retrospective chart reviews, 4 mixed-methods, 4 qualitative (including 2 case studies) | Adults aged over 18 with advanced cancer receiving telehealth interventions |  |  | Template for Intervention Description and Replication (TIDieR) checklist |  |
| Miranda et al., 2019 ^68^ | Systematic review | Up to 2018 | n= 8 | n= not provided | Specialist palliative care/Non-specialist palliative care | Symptom, Functional status, behavioural symptoms, Place of death, satisfaction | Yes | AMSTAR - 2 rating: Low |
| Belgium and United Kingdom |  |  | Retrospective case-control (n=2); retrospective cross-sectional (n=1); RCT (n=4); and an unclear design (n=1) | Patients with dementia, living at home, and receiving palliative care at home. |  |  | Quality Assessment Tool for Quantitative Studies’ developed by Effective Public Health Practice Project |  |
| Mojtahedi and Shen, 2023 ^69^ | Scoping review | Not provided | n= 12 | n= 21014 | A palliative care team delivering home hospice care through the pandemic | Perspectives of patients/caregivers/healthcare providers on home palliative care during the COVID-19 pandemic | No | No adherence to PRISMA-ScR or equivalent |
| United States of America |  |  | Qualitative (n=10, of which 7 are survey based, 1 was telephone SSI, 2 were case-studies); Quantitative through medical record data(n=2) | Patients and caregivers receiving home palliative care or healthcare providers delivering palliative care during the COVID 19 pandemic. Home care was defined as being received at home either face to face or through telehealth. |  |  |  |  |
| Nordly et al., 2016 ^90^ | Systematic review | 2000-2015 | n= 8 | n= not provided | Specialist Palliative care delivered primarily in the home by a team with a least a doctor or nurse on the team | Place of death, survival, quality of life, performance status, pain and dyspnoea | Yes | AMSTAR - 2 rating: Critically low |
| Denmark |  |  | Longitudinal Observational (n=2); Observational cross sectional (n=4); Interventional before and after n=2 | Patients receiving home-based specialist palliative care with advanced cancer. Advanced cancer defined and incurable or metastatic |  |  | Not stated |  |
| O’Connor et al., 2022 ^70^ | Scoping review | 1995 -2020 | n= 18 | n= not provided | Community based palliative care services for people with dementia and their carers | Caregivers: Pain, QoL, satisfaction, ACP, caregiver burden, place of death, functional status, behavioural symptoms, burden, Depression, functional and cognitive status, hospital admissions, number of referrals to hospice palliative care team | Yes | No adherence to PRISMA-ScR; Utilised Levac et al., (2010) methodology for Scoping reviews |
| Ireland |  |  | Randomised control trials (RCTs) (n =12); retrospective cohort (n =6); prospective cohort (n =3); cross-sectional (n =4); pre-post (n =3); case-control (n =2) | People with moderate to severe dementia of any type, or carers of people with dementia, living in the community in their family homes, or a nursing home setting. |  |  | Guidance by Scoping studies: advancing the methodology. Implementation science |  |
| Patton et al., 2021 ^71^ | Systematic review | 2019-2021 | n= 10 | n= not provided | Specialist palliative care & outpatient consultations | Pain, Other symptoms, Quality of life, Satisfaction, Referral to other services, Satisfaction, Burden/morbidity, Home death rates, Health service use and costs, Increased adherence to guidelines, Prescribing rationale Health care/voluntary sector costs | Yes | AMSTAR - 2 rating: Critically Low |
| United Kingdom |  |  | 6 retrospectives observational, 3 longitudinal, 1 prospective pilot phase II | Adult palliative care patients suffering from cancer. |  |  | Evidence-based librarian (EBL) critical appraisal checklist |  |
| Peeler et al., 2023 ^72^ | Scoping review | Up to 2022 | n= 18 | n= 2508 | Community engaged end of life care | Practical needs (physical, education, psychological, spiritual and social support); Personal growth (knowledge, skills, and attitudes about death and dying; personal reflection and confidence); Community capacity (developing community activists, embedding sustainable change) | No | Adhered to the Preferred Reporting Items for Systematic Reviews and Meta-analyses Extension for Scoping Reviews (PRISMA-ScR) checklist |
| United Kingdom |  |  | Randomized control trials (n = 8), six qualitative only, two quasi-experimental, and two pre-post designs | Adults aged 18 and over within the last year of life and their carers, with interventions delivered in the community |  |  |  |  |
| Peerboom et al., 2023 ^73^ | Scoping review | Inception to August 20, 2022 | n=9 | n=2280 | End-of-life communication | Perspectives of the nursing staff, family caregiver, and the older person on end-of life communication | No | Adhered to the Preferred Reporting Items for Systematic Reviews and Meta-analyses Extension for Scoping Reviews (PRISMA-ScR) checklist |
|  |  |  | Qualitative (n=6), Quantitative (n=3) | Nursing staff (i.e., care assistants, certified nursing assistants, licensed vocational nurses, registered nurses, clinical nurse specialists, nurse practitioners), or (family caregivers of) older people in the hospital, nursing home or home care setting. |  |  |  |  |
| The Netherlands |  |  |  |  |  |  |  |  |
| Pinto et al., 2024 ^74^ | Umbrella review | Inception to October 11, 2022 | n=15 | n=141,159 | End of life care | Preferences about place of end-of-life care and death of patients with life-threatening illnesses and their families. | Yes | AMSTAR - 2 rating: Moderate |
|  |  |  | Six reviews were quantitative (two with meta-analysis), three were qualitative, and six were mixed-methods (one with meta-analysis). | Patients diagnosed with life-threatening illnesses and/or their family members (of any age, gender and race/ethnicity). |  |  | JBI Critical Appraisal Checklist for Systematic Reviews and Research Syntheses. |  |
| Portugal, United Kingdom |  |  |  |  |  |  |  |  |
| Rabow et al., 2013 ^91^ | Narrative review | Not provided | n= 4 | n = not provided | Outpatient non-hospice palliative care | Quality of life, symptom burden, spiritual wellbeing, sleep quality, satisfaction with care, survival and resource use | No | SANRA Score= 9 |
| United States of America |  |  | Prospective randomised controlled trial (3) and prospective cluster randomised controlled trial (1) | Patients with advanced illness (including cancer and non-cancer conditions |  |  |  |  |
| Sánchez-Cárdenas et al., 2022 ^75^ | Systematic review | 2010-2022 | n= 14 | n= 5485 | Telemedicine for patients cared for by primary care teams, those located in rural areas or challenges with travelling, | Symptom management, caregiver support, psychosocial support, pharmacological monitoring, patient follow up, health education, QoL | Yes | AMSTAR - 2 rating: Low |
| Columbia |  |  | Observational (6), experimental (6) and qualitative (1) | Palliative care for advanced cancer patients with difficulties in accessing standard care (i.e. those living rurally) |  |  | The Newcastle-Ottawa Scale and Standards for Reporting Qualitative Research (SRQR) |  |
| Sani et al., 2024 ^76^ | Systematic review | Inception to October 30, 2023 | n=7 | n=376 | Home-based interventions, such as self-rehabilitation (rehabilitation exercises), educational intervention, structured planned home visits, and pulmonary rehabilitation. | Quality of life (QoL), adherence to treatment, fatigue, bimanual and related activities | Yes | AMSTAR - 2 rating: Moderate |
|  |  |  | RCTs (n=4) and Quasi-experimental studies (n=3) | Patients with different terminal illnesses |  |  | Revised Cochrane risk of bias tool for randomized studies (ROB2) |  |
| Nigeria |  |  |  |  |  |  |  |  |
| Santos et al., 2022 ^77^ | Systematic review | 2015-2019 | n= 7 | n= not provided | Not specified beyond palliative care | Caregiver satisfaction and opinions, cost, place of death | Yes | AMSTAR - 2 rating: Critically low |
| Brazil |  |  | Not stated: objectives and results only | Not stated beyond "elderly" |  |  | Tool unreported |  |
| Santos et al., 2023 ^78^ | Systematic review | 2012– November 2022 | n=8 | n=763 | Palliative care interventions | Quality of life, Symptom burden and control, Anxiety and depression, Advanced care planning, Hospitalisations, and Survival. | Yes | AMSTAR - 2 rating: Low |
|  |  |  | RCT (n=7) and Cluster-controlled trial (n=1) | Individuals diagnosed with chronic non-malignant respiratory diseases |  |  | Revised Cochrane risk of bias tool for randomized studies (ROB2) |  |
| Portugal |  |  |  |  |  |  |  |  |
| Sarmento et al., 2017 ^92^ | Systematic review | 2000 onward | n= 19 | n= 814 | Home-based palliative care | Experiences and components of home palliative care | Yes | AMSTAR - 2 rating: Low |
| United Kingdom, Portugal and the Netherlands |  |  | Interviews (n=14); case study (n=1); surveys (n=3); analytic expansion of studies with statement analysis and theory synthesis (n=1) | Adult patients (aged 18+) with a life-limiting diagnosis and palliative care needs and/or their family caregivers being cared for at home |  |  | A modified version of the CASP (Critical Appraisal Skills Programme) criteria |  |
| Seica Cardoso et al., 2023 ^79^ | Systematic review | inception until October 2022 | n=4 | n=268 | Non-pharmacological interventions | Patients’ quality of life | Yes | AMSTAR - 2 rating: High |
|  |  |  | RCT (n=4) | Patients with palliative care needs |  |  | Cochrane risk-­of-­bias (RoB) V.2.0 |  |
| Portugal, United Kingdom |  |  |  |  |  |  |  |  |
| Shepperd et al., 2021 ^80^ | Systematic review | Inception to 2020 | n= 4 | n= not provided | Home-based end-of-life care | Place of death, hospital admissions, patient satisfaction, caregiver satisfaction, Cost | Yes | AMSTAR - 2 rating: High |
| United Kingdom |  |  | Randomised control trials | People aged 18 years and over who were receiving end-of-life care at home, at the end of life and required terminal care. |  |  | Cochrane 'Risk of bias' criteria |  |
| Spencer et al., 2024 ^81^ | Rapid review | January 2003 to October 2023 | n=56 | n = not provided | Hospital based model of care, Home/community- based model of care, Hospice based model of care, Mixed models of care | Cost-effectiveness, cost of palliative care or end of life care. | Yes | AMSTAR - 2 rating: Critically low |
|  |  |  | Systematic reviews (n=8), Primary studies (n=48) | Children and adults receiving palliative or end of life care |  |  | Joanna Briggs Institute (JBI) economic evaluations checklist |  |
| United Kingdom |  |  |  |  |  |  |  |  |
| Ventura et al., 2014 ^93^ | Systematic review | 1975 to 2012 | n= 15 | n= 728 | No intervention of interest: qualitative review of unmet needs identified in relevant literature | Unmet needs | Yes | AMSTAR - 2 rating: Critically low |
| Australia |  |  | Qualitative interviews (n=10); quantitative cross-sectional (n=2); mixed qual/quant interviews (n=2); and quantitative questionnaire (n=1) | Adult palliative care patients and/or their informal caregivers |  |  | The Standard Quality Assessment Criteria for Evaluating Primary Research Papers from a Variety of Fields |  |
| Vernon et al., 2022 ^82^ | Systematic review | Up to 2021 | n= 61 | n= not provided | Community-based palliative care | Place of death, hospitalisation, Emergency Department (ED) visits, QoL, cost | Yes | AMSTAR - 2 rating: Low |
| United States of America |  |  | Mixed methods (n=18) Quantitative (n=50) | Those in receipt of community-based palliative care regardless of patient characteristics |  |  | Mixed Methods Appraisal Tool version 2018 |  |
| Walshe and Luker, 2010 ^96^ | Realist review | 1990-2009 | n= 46 | Not stated | District nursing, or equivalent, role in home-based palliative care | Value of in-home nursing; resource availability; DN role definition; resource allocation; symptom management; interprofessional communication; patient and informal caregiver satisfaction; patient and caregiver education; and caregiver resources | Yes | AMSTAR - 2 rating: Critically low |
| United Kingdom |  |  | Qualitative data from interviews n=33; Observational n=6; Questionnaires n=8. |  |  |  | Using the guiding principle for quality appraisal in realist reviews (Pawson, 2006) |  |
| Wicaksono et al., 2024 ^83^ | Scoping review | Inception to April 18, 2023 | n=24 | n = not provided | Home based palliative care | Perspectives of the family caregiver | No | Adhered to the Preferred Reporting Items for Systematic Reviews and Meta-analyses Extension for Scoping Reviews (PRISMA-ScR) checklist |
|  |  |  | Qualitative research methods 23 studies and One mixed-method study | Family caregivers of people receiving home palliative care |  |  |  |  |
| The Netherlands, Indonesia |  |  |  |  |  |  |  |  |
| Zheng et al., 2016 ^94^ | Systematic review | 2003-2015 | n= 9 | n= not provided | Telehealth interventions for caregivers of patients receiving palliative care. | Quality of life, anxiety, | Yes | AMSTAR - 2 rating: Critically low |
| United States of America |  |  | Prospective exploratory cohort study, sequential mixed methods, two phase design, two group nonrandomised design, pooled analysis of two randomised trials., mixed methods analysis, pilot study, mixed methods case study, randomized noninferiority trial with two groups, single group feasibility study | Patients receiving palliative or end-of-life care for a serious condition (i.e., advanced disease, end-stage disease). Caregivers were adults >18yr, relationships to patients included spouses/partners, parents, Children, siblings, grandchildren, daughter-in-law. |  |  | Cochrane Collaboration’s tool for assessing risk of bias |  |
| Zimbroff et al., 2021 ^84^ | Systematic review | Up to 2019 | n= 16 | n= not provided | Home based palliative care interventions | Cost, health service utilisation (including inpatient admissions, hospital days, and skilled nursing facility use), quality of care, and patient/caregiver satisfaction with care, community survival, and mortality rate. | Yes | AMSTAR - 2 rating: Critically low |
| United States of America |  |  | Case study (n=2); programme description (n=1); retrospective chart review (n=4); case-cohort study (n=1); commentary (n=1); survey (n=3); case-control study (n=1); retrospective cross-sectional study (n=1); cost projection (n=1); and expert perspectives (n=1) | Homebound Medicaid beneficiaries aged 18 and older in receipt of home-based primary care (HBPC) or home-based palliative care (HBPalC) |  |  | Newcastle-Ottawa Scale for quality assessment |  |
